# Multiscale integration of human and single-cell variations reveals unadjuvanted vaccine high responders are naturally adjuvanted

**DOI:** 10.1101/2023.03.20.23287474

**Authors:** Matthew P. Mulè, Andrew J. Martins, Foo Cheung, Rohit Farmer, Brian Sellers, Juan A. Quiel, Arjun Jain, Yuri Kotliarov, Neha Bansal, Jinguo Chen, Pamela L. Schwartzberg, John S. Tsang

**Author notes:** Correspondence (JST).

## Abstract

Advances in multimodal single cell analysis can empower high-resolution dissection of human vaccination responses. The resulting data capture multiple layers of biological variations, including molecular and cellular states, vaccine formulations, inter- and intra-subject differences, and responses unfolding over time. Transforming such data into biological insight remains a major challenge. Here we present a systematic framework applied to multimodal single cell data obtained before and after influenza vaccination without adjuvants or pandemic H5N1 vaccination with the AS03 adjuvant. Our approach pinpoints responses shared across or unique to specific cell types and identifies adjuvant specific signatures, including pro-survival transcriptional states in B lymphocytes that emerged one day after vaccination. We also reveal that high antibody responders to the unadjuvanted vaccine have a distinct baseline involving a rewired network of cell type specific transcriptional states. Remarkably, the status of certain innate immune cells in this network in high responders of the unadjuvanted vaccine appear “naturally adjuvanted”: they resemble phenotypes induced early in the same cells only by vaccination with AS03. Furthermore, these cell subsets have elevated frequency in the blood at baseline and increased cell-intrinsic phospho-signaling responses after LPS stimulation *ex vivo* in high compared to low responders. Our findings identify how variation in the status of multiple immune cell types at baseline may drive robust differences in innate and adaptive responses to vaccination and thus open new avenues for vaccine development and immune response engineering in humans.

## Introduction

Human immune systems exhibit substantial person-to-person variation^1–4^. Population variations in immune response outcomes to the same perturbation, such as antibody responses to vaccination, can be linked to cellular and molecular immune system components using top-down systems biology approaches^4,5^. Such studies have used unbiased immune profiling to identify signatures of response to perturbations and predictors of outcomes such as antibody response to vaccination^6–14^, uncovering contributions from intrinsic factors, such as genetics^15^, age^16,17^, and sex^18^. Furthermore, accumulating evidence from these studies supports the hypothesis that immune system status prior to a perturbation can predict and potentially influence both response quality and quantity^6,16,19–23^. For example, we identified transcriptome signatures reflective of an immune system “set point” predictive of higher antibody response following vaccination in healthy individuals^22^; the same signature when evaluated during relative clinical quiescence was also linked to increased plasma cell related transcriptomic activity during disease flares in lupus patients. More recently, blood transcriptome profiling studies identified prognostic signatures in healthy children at risk of type 1 diabetes prior to development and onset of the disease^24^, and at baseline in cancer patients prior to immunotherapy induced autoimmunity^25,26^.

While the biomarker signatures identified thus far are informative, technological limitations hinder a high-resolution and holistic view of immune cell processes that underlie baseline set points that predict and potentially determine optimal responses^27,28^. Bulk blood transcriptomic profiles are confounded by substantial inter-individual variations in circulating immune cell subset frequency^6,29,30^, while protein based phenotypes measured using cytometry alone often cannot assess internal cell states such as those captured by transcriptomics. Single cell transcriptomics can better resolve cell states but interpretation remains challenging when measuring chromatin accessibility or mRNA alone without utilizing, for example, existing knowledge cataloging immune cell types and subsets using surface protein markers^6,29–31^. Multi-modal single cell transcriptome and protein profiling methods such as CITE-seq^32^ are promising for unifying these modalities; however, the integrative analysis of timed perturbation responses including the decomposition of meaningful biological variations spanning different size scales from individual human subjects to cell types and single cells remains a major challenge.

In this work, we developed a multilevel modeling framework to integrate human population, temporal, and single cell variations. We applied this framework to extract vaccine response kinetics and cell states, and attributed cell type specific transcriptomic variations to age, sex, subject, perturbation, and time. Using CITE-seq^32^, we profiled PBMCs from 26 subjects before and after vaccination with two different pandemic influenza vaccines. Individuals were nested into three groups: those with 1) high or 2) low antibody responses to an unadjuvanted influenza vaccine and 3) individuals vaccinated with an AS03 adjuvanted vaccine against H5N1 influenza. We further revealed previously unknown, cell type specific phenotypes specifically induced by AS03^33^. In addition, we unbiasedly defined the landscape of baseline immune phenotypes linked to high antibody responses, demonstrating that these do not merely reflect the phenotypes of a single cell type but instead capture an extensive correlated set of phenotypes across different cell types. Furthermore, by comparing the baseline (prevaccination) cell type specific predictors of unadjuvanted vaccine responses with phenotypes induced specifically by the unadjuvanted influenza vaccine, the COVID-19 mRNA vaccine, and the AS03 adjuvanted H5N1 vaccine revealed that high responders to the unadjuvanted vaccine were “naturally adjuvanted” at baseline. This concept was further buttressed by data from phosphoprotein signaling responses to *ex vivo* cell stimulation. Our integrative approach paves the way for multiscale analysis of timed perturbation studies using multimodal single cell data in humans. Furthermore, our findings suggest cell type specific targets of immune response engineering and vaccine development.

## Results

### Multimodal single cell profiling to assess human response variations to timed vaccine perturbations

To generate a multimodal single cell data set that captured biological variations spanning molecular and cellular states, vaccine formulations, inter- and intra-subject differences, and response kinetics, we assessed 52 PBMC samples from 26 donors pre- and post-vaccination using CITE-seq (Figure 1a). Subjects received either the 2009 seasonal and pandemic type A strain vaccine combination, or an H5N1 avian influenza strain formulated with oil in emulsion adjuvant AS03^6,34^. For the AS03 group, we focused on the baseline and innate response (day 1) time-points since AS03 is known to elicit a strong early response^35,36^. For the unadjuvanted seasonal influenza vaccine, twenty subjects with high (n=10) and low (n=10) antibody responses were selected from our cohort of 63 individuals that we previously profiled and stratified into high, mid, and low responders based on antibody titer fold change adjusted for age, sex ethnicity and pre-existing immunity^6,22^. These 20 individuals were profiled at baseline and select subsets of individuals on day 1 or 7 post vaccination to assess the innate and adaptive cellular responses (**Figure 1a**). We analyzed sources of technical noise in CITE-seq surface protein expression data by using our recently developed normalization method called dsb^37^, then assessed the robustness of CITE-seq to recover and unify known cell surface and transcriptome phenotypes. For example, both activated B cells and plasmablasts could be distinguished based on the expression of CITE-seq surface protein markers CD19, CD71, CD20, and CD38. We further confirmed that the gated cells exhibit transcriptional signatures^38^ derived previously from these cell subsets after FACS-sorting (**Figure S1a**).

**Figure 1.**
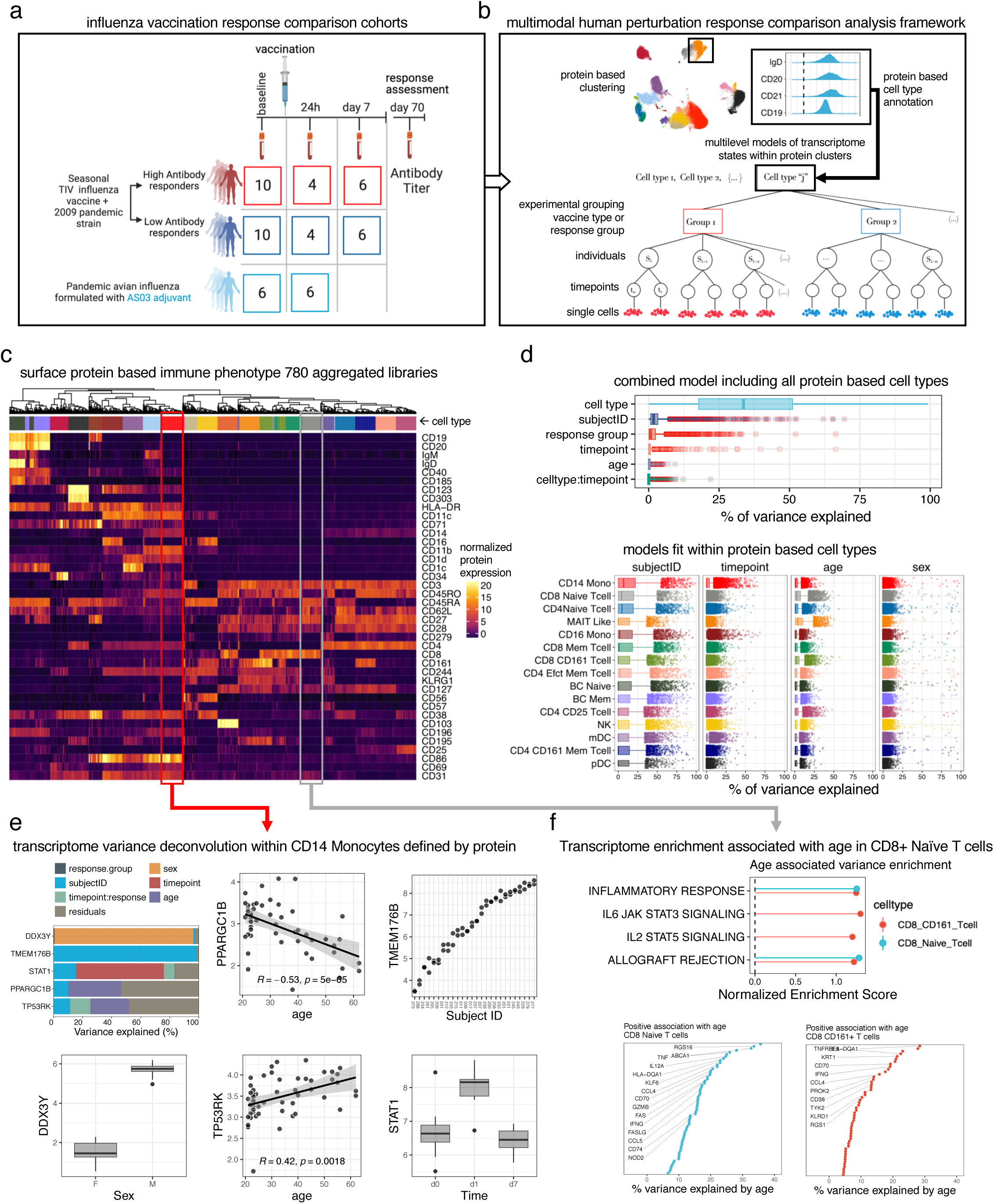
Multimodal single cell portraits of human vaccination response through within cluster mixed models comparing vaccination effects over time. **a.** Human vaccination response study outline; CITE-seq data was generated from n=52 PBMC matched pre- and post-vaccination PBMC samples from n=26 subjects including 2 response groups and two vaccine formulations. Numbers in the boxes indicates the number of samples run with CITE-seq. 10 high and 10 low responders from the 2009 TIV + pandemic H1N1 influenza vaccination without adjuvant were profiled with a subset of 8 and 12 subjects split evenly between high and low responders profiled on day 1 and 7 respectively. 6 subjects vaccinated with a pandemic H5N1 avian influenza vaccine formulated with adjuvant AS03 were profiled at baseline and day 1 post vaccination. **b.** The hierarchical structure of the data for a single cluster is shown to motivate necessity of multilevel modeling approach for transcriptome analysis. Clusters are based on surface protein (select proteins from naïve B cell cluster shown); within each cluster modeled with weighted mixed effects models clusters are represented by cells from PBMC samples indexed by individual, timepoint and different response groups (high and low responders) and vaccine group (unadjuvanted vs adjuvanted). **c.** For each of 780 samples aggregated by protein based cell type and individual x timepoint, the median dsb normalized protein expression in each cell type is shown–colors of cell types are the same as shown in d. **d.** Top: the fraction of variance explained in a multivariate model across libraries aggregated by cell type, individual and timepoint; bottom: as in the top panel, but here with models fit within each protein based cell type, i.e. within colored columns of c. **e.** Variance fractions for an example group of 5 genes from the multivariate mixed model fit within CD14 monocytes with additional visualizations of gene expression (y axis) vs the experimental factor (x axis) explaining maximal variance for the 5 genes. **f.** Top: enrichment of pathways in the MsidDB Hallmark gene sets based on genes ranked by their variance explained by age; subset of genes with positive association with age in CD8 naïve and CD161+ T cell clusters; bottom: select genes positively associated with age within the two cell types.

### Transcriptome variation decomposition into surface protein-based cell type, individual, age, sex, and vaccination effects

Cells clustered using the 82 surface proteins were enriched for known immune phenotypes **(Figure S1 c, d)**. Cells from individual subjects at different timepoints were represented in a majority of cell clusters **(Figure S1e,f)**. Some cell clusters were dominated by cells from two to three subjects (e.g., NKT and CD57+ CD4 T cells); this likely reflects individualistic phenotypes as the samples from different timepoints from the same individuals were also present in the same clusters, suggesting that these phenotypes represent temporally stable, within-individual variations^6^ **(Figure S1f)**.

Instead of analyzing one variable at a time, we next deconstructed the transcriptional variation of each gene into that attributable to cell types, individuals, intrinsic factors (age, sex), and vaccination responses (**Figure 1b**) using multivariate mixed effects models. For each gene, these models quantify contributions of biological factors (such as cell type or subject effects) toward observed expression variation, including adjusting for dependency among repeated measures from the same individuals (see Methods). Models were first fit to each transcript across 780 transcriptome (“pseudobulk”) libraries indexed by cell type, individual, and timepoint **(Figure 1c, columns)**. Variance patterns for each gene in every cell type are provided in Supplemental Tables 3 and 4. This analysis revealed that cell type explained more than 30% of the variation across the transcriptome (range 0-100%; **Figure 1d, top**); this observation is consistent with the fact that different cell types have distinct transcriptome profiles^39,40^. To identify cell type intrinsic and vaccination effects independent of differences among cell types, we next fit models within the cell subsets defined by surface proteins **(Figure 1d, bottom)**. For example, this analysis revealed factors contributing to the extensive differences among CD14+ classical monocytes; 5 example genes are shown in (**Figure 1e**). As expected, sex almost completely explained the variation in the expression of a Y-linked gene (DDX3Y). A transcription factor genetically linked to rheumatological pathology^41^ (PPARGC1B) and an apoptosis regulator (TP53RK) were negatively and positively associated with age, respectively. Overall, our approach identified substantial between-subject variations for many genes (**Figure 1d**, see “SubjectID”). For example, inter-subject differences accounted for nearly 100% of expression variation in TMEM176B, an inflammasome signaling regulator^42^, suggesting that inflammasome function could have substantial individuality in the human population. Temporal variation (e.g., differences relative to baseline following vaccination) accounted for more than 50% of the expression differences in STAT1; a separate differential expression model revealed that vaccination induced expression of this gene within monocytes a day after vaccination (see below). Age was also a major contributor, particularly in genes within the CD8 naïve and CD8+ CD161+ T cells relative to other cell types; inflammatory processes were specifically enriched among genes positively correlated with age (**Figure 1f**), consistent with sterile inflammation linked to aging^43^ or “inflammaging”.

Thus, our approach provides a global view of the extent by which different biological factors contribute to gene expression variation.

### Single cell deconvolution of the early response to unadjuvanted influenza vaccination reveals both cell type-specific and -agnostic patterns

Given that most of the known transcriptional response signatures of vaccination were derived using whole blood/PBMC profiling, we next assessed time-associated changes from our mixed effects models to identify cell type specific responses elicited by unadjuvanted vaccination on days 1 and 7 (after modeling between individual variation and adjusting for age, sex, baseline antibody titers, and other technical factors - see Methods). Gene set enrichment analysis revealed that day 7 responses comprised naïve B cell and CD4+ memory T cell activation and metabolic processes; however, some are not significant after FDR correction and these effects were generally weaker than early response effects described below **(Figure S2a,b).** Changes in circulating plasmablast frequencies were thought to drive whole blood transcriptome signatures (typically measured on day 7-12 post vaccination) predictive of antibody response to multiple vaccines^6–8,44^. Indeed, here plasmablasts had the most elevated signature score (i.e., average expression of genes we compiled based on previous day-7 bulk transcriptome signatures predictive of antibody responses) in our day 7 vs. day 0 comparison relative to other cell subsets **(Figure S2c).** B cell maturation antigen (BCMA) receptor (TNFRSF17) had the highest fold change in both bulk microarray and “pseudobulk” CITE-seq data **(Figure S2d).** Deconvolution of the CITE-seq sequencing reads to each cell type revealed that nearly all the TNFRSF17 counts (see Methods) were derived from the day-7 CD38high CD20-plasmablast cells and not from naïve or memory B cell subsets **(Figure S2e).**

Unadjuvanted influenza vaccination response studies consistently report interferon stimulated gene expression (ISG) detected early (1-3 days) post vaccination in bulk blood transcriptomic data. Furthermore, elevation of ISG and antigen presentation genes on day 1 has been found to correlate with higher antibody response^11^, although the cellular origins of these responses were not fully resolved. Based on microarray profiling of sorted cell subsets, early reports suggested that this signal originated primarily from DCs on day 3^45^ or monocyte/granulocytes on day 1^13^. Here, unbiased CITE-seq assessment using curated gene sets, including influenza vaccine response signatures obtained from the literature that were derived from bulk transcriptomic data (See **Supplementary Table 1**), identified three broad patterns of responses 24 hours following vaccination. The first pattern was characterized by genes downstream of type I and type II interferon signaling pathways that are shared across cell types (Figure 2a). 46 shared, “core genes” were collectively induced in at least 5 cell types (Figure S2f), including the transcription factors IRF1 (notably, induced across 15 cell types), STAT1, IRF7, and IRF9. Also included were pattern recognition receptor (PRR) genes IFITM1 and IFITM3, inhibitors of vial transcription GBP1^46^ and ISG15^47^, and antigen presentation genes TAP1, and PSMB9 (Figure S2f). The second pattern encompassed responses unique to classical and nonclassical monocytes, such as adhesion molecule ICAM1, JAK2, antigen presentation / HLA genes, and inhibitors of viral replication OAS3^48^, and ISG20^49^. The third pattern pointed to more individual cell-type specific responses (Figure S2g), notably, inflammatory processes induced within classical monocytes. The “reactome interferon signaling” genes (**Figure 2a**) captured all three response patterns, with 10-15 shared ISGs across multiple cell subsets, a specific set of ISGs shared by classical and non-classical monocytes, and a set of classical monocyte specific genes (**Figure 2b**). The expression of these genes in classical monocytes alone clustered samples by time relative to vaccination, suggesting that they were induced in a coordinated manner across individuals after vaccination (**Figure 2c**).

**Figure 2.**
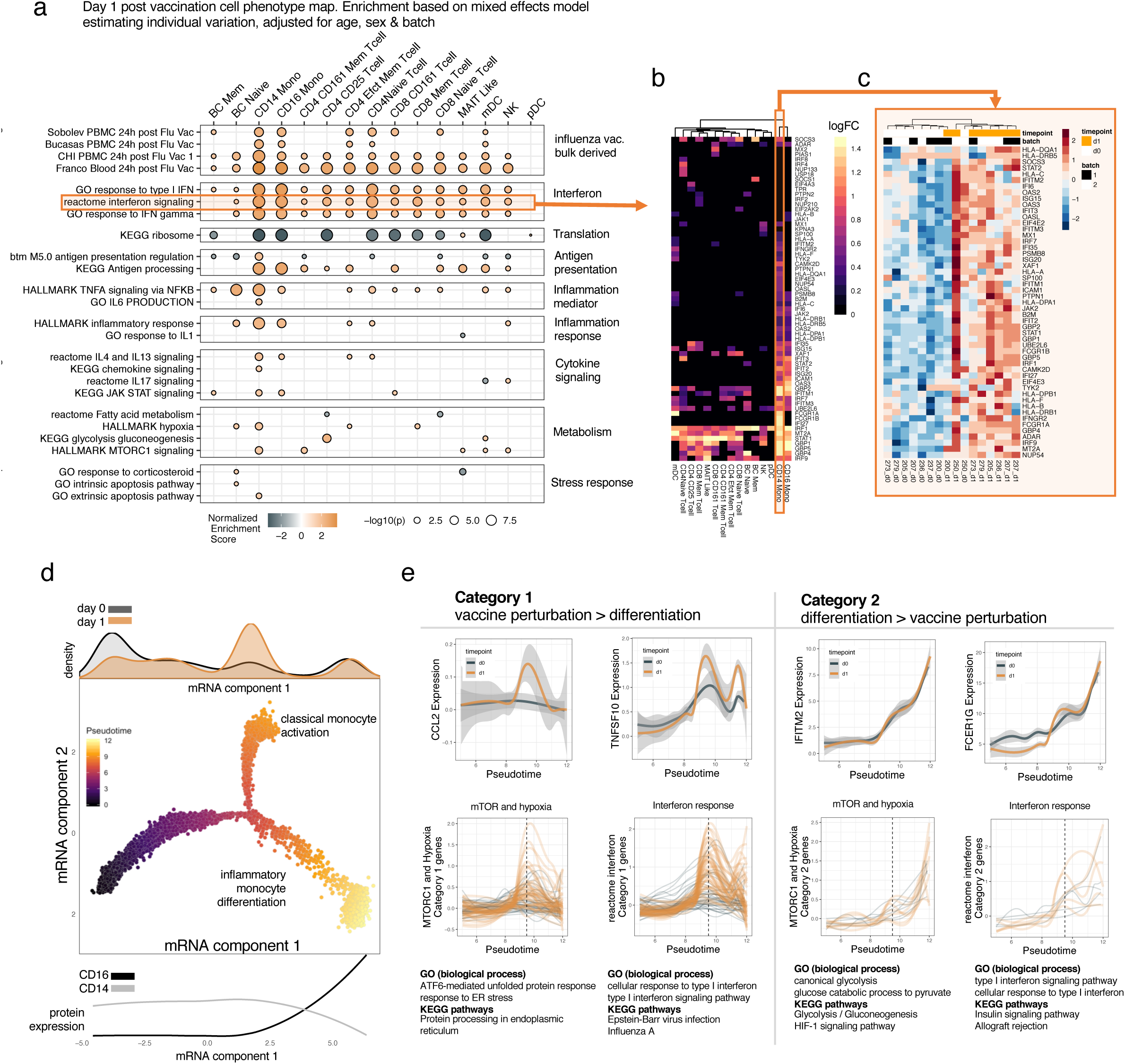
Top down and bottom up deconstruction of transcriptome perturbations induced day 1 post vaccination with seasonal TIV + 2009 pandemic strain vaccine. **a.** Day 1 post vaccination transcriptional response within protein-based cell types. Gene set enrichment (orange = positive enrichment/upregulation, black= negative enrichment/ downregulation) of modules based on genes ranked by pseudobulk weighted linear mixed effects model baseline vs day 1 effect size. The broad category of each curated module / pathway is labeled on the right margin; see supplemental table 1. **b.** Leading edge genes from the reactome interferon signaling module; cell types shown with enrichment at adjusted p value < 0.05. **c.** Log counts per million of aggregated data for each subject within CD14 monocytes defined by protein of leading edge genes from the “interferon signaling” module CD14 monocyte day 1 enrichment demonstrate a coordinated post vaccination across individuals (hierarchically clustered genes and samples). **d.** DDR-tree algorithm constructed with baseline and day 1 post vaccination cells. Component 1 and component 2 are latent space embeddings based on mRNA only for single monocytes as determined by the DDR-tree algorithm. Each point is a single cell and is labeled by pseudotime as calculated by monocle. The timepoint relative to vaccination of each cell along mRNA trajectory component 1 is highlighted in the top marginal histogram; cells are colored by inferred pseudotime. Three branches from left to right are enriched for resting classical monocytes, activated classical monocytes from post vaccination, and nonclassical monocytes. Cells progressively downregulate CD14 and upregulate CD16 protein level along the rightmost branch; protein data shown in the bottom margin basis spline fit to dsb normalized protein level for CD14 and CD16 (protein levels were not used to construct the trajectory). **e.** Gene expression of select leading edge genes from enrichments in CD14 monocytes based on branch-dependent differential expression show two broad patterns. Pattern 1 genes are perturbed by vaccination with highest expression in post vaccination classical monocytes – dashed line at pseudotime value of 9.5 represents the peak of activation. Pattern 2 genes continuously increase across pseudotime and have highest expression in CD16+ CD14-non-classical monocytes. The top row shows example genes from each category. The bottom row shows the subset of genes falling into each category from the combined hallmark MTORC1 signaling/Hypoxia pathways and reactome interferon signaling pathways. Below each category / pathway, enrichment of gene ontology (GO) biological process and KEGG pathways for the subset of genes from each pathway and category.

Genes driving the classical monocyte “IL6 production” pathway reflected early initiators of inflammation MYD88, DDX-58 (RIG-I), TNF and TRAF6. Inflammatory processes were further implicated by monocyte specific expression of IL-15, and chemokine CCL2^50^ (Figure S2g). Classical monocytes were also enriched for hypoxia and mTORC1 signaling pathways (**Figure 2a**). While natural influenza infection can activate and subvert mTOR signaling to support viral replication^51^, this signal following inactivated vaccination was more likely to reflect the role of mTOR in inflammation^52^. The genes driving this enrichment signal (“leading-edge genes”) suggested that mTOR induced glycolytic metabolism might be involved: this process is known to be induced after VZV vaccination^53^ and is linked to non-specific innate memory in monocytes^54^. mTOR enrichment within CD25+ CD4 effector T cells, MAIT-like cells, mDCs and NK cells may have been intrinsically induced by TIV or by monocyte specific expression of IL-15 (Figure S2g), a cytokine that can activate mTOR in human NK cells^55^. These cell-specific and shared unadjuvanted vaccine response perturbations and driver genes are provided in **Supplementary Table 2**.

We next explored how time associated response signatures from our statistical models could be coupled to “bottom up” single cell computational reconstructions of transcriptional dynamics induced by vaccination. By using single monocytes from both days 0 and 1 samples, we derived a pseudotime, tree-based latent cell-phenotype space via a “reversed graph embedding” algorithm^56,57^ (**Figure. 2d**). CD14 and CD16 surface protein expression patterns allowed identification of cell subset enrichment at the ends of the three tree branches (**Figure 2d**, bottom margin): pre-vaccination classical monocytes along the left branch, their day 1 counterparts in the top branch, and the non-classical monocytes from both before and after vaccination enriched in the right branch. Integrating the monocyte specific vaccination response phenotypes from above (Figure 2a) with this latent space visualization identified two categories of genes based on branch-dependent differential expression (see Methods). Category 1 genes mainly reflected vaccine perturbation effects within either CD14 monocytes alone (e.g., CCL2) or both within CD14 and CD16 monocytes (e.g., TNFSF10), whereas category 2 genes (e.g., IFITM2, FCERG1) captured differences and potential differentiation between classical and non-classical monocytes; these genes continuously increased across the spectrum of pseudotime with the highest expression in nonclassical monocytes (**Figure 2e**, top row). This analysis also revealed that IFN response genes in Figure 2c mostly belonged to category 1 (more than 40 genes) except for 5 genes, PTPN1, IFITM2, IFITM3, HLA-C and EIF4E2 which belonged to category 2. The mTOR and hypoxia pathway genes followed a similar pattern, though notably the genes falling in category 2 were more enriched for glycolysis than those in category 1, which were more enriched for ER stress (**Figure 2e**, bottom). These results illustrate how integrating effects associated with day 1 changes following vaccination (“real time”) and single-cell latent space/pseudotime reconstruction can highlight interwoven cellular activation and differentiation processes and reveal finer shades of phenotypic variation in response to vaccination.

### The AS03 adjuvant induces unique myeloid innate-sensing and B-cell anti-apoptosis enhancement signatures compared to unadjuvanted vaccination

We next examined early response (day 1) variations attributable to the vaccine adjuvant AS03. AS03 is known to elicit both higher level and diversity of anti-influenza antibodies compared to unadjuvanted vaccines, even when formulated with a low antigen dose^33^. Previous studies of transcriptional responses to AS03 adjuvanted vaccines revealed strong early induction of ISGs in innate immune cells^33,35,36,58^ when comparing against a low-dose antigen control formulated with PBS. Here we applied a statistical contrast defining the difference in the day 1 responses (relative to baseline) between the AS03 adjuvanted vaccine versus the unadjuvanted vaccine described above. We then validated these signatures using an independent data set from profiling FACS-sorted immune cells (e.g., total B and T cells) from subjects receiving the same vaccine formulated with AS03 versus PBS^58^ **(Figure 3a, Figure S3a)**. We first noticed positive enrichment of several pathways related to surface receptors in monocytes and mDCs **(Figure 3b, red);** these were highly concordant with data from similar innate cell subsets in the validation cohort (**Figure 3b**, **light blue).** The leading-edge genes driving these enrichments include immune receptors recognizing different classes of pathogens (beyond just the receptors recognizing specific molecular patterns in the vaccine), thus suggesting expansive upregulation of receptors to increase the capacity of cells to sense environmental signals. For example, Toll Like Receptors (TLRs) recognizing both bacterial and viral molecular patterns TLR1, TLR4, TLR5, and TLR8 were among the leading-edge genes in the CD14 monocyte module “M16”, as was FPR2, which is known to induce immune cell chemotaxis in response to bacterial metabolites^59^. Examination of genes with strong AS03-specific effects beyond genes in these specific sensing pathways identified additional PRRs in monocytes, for example c-GAS, a cytosolic DNA sensor that activates antiviral response via STING^60^ (**Figure 3c**). Within mDCs, day 1 enrichment of the “rhodopsin like receptors” module was driven by genes related to inflammatory chemotaxis such as FPR1^59^ and CCR1^61^ and P2RY13, an ADP sensor active during inflammation^62^, which were induced to a greater degree by AS03. As with CD14+ monocytes, mDCs also had evidence of AS03 specific upregulation of TLR4, the PRR for bacterial lipopolysaccharide^63^ (**Figs. 3c-d**).

**Figure 3.**
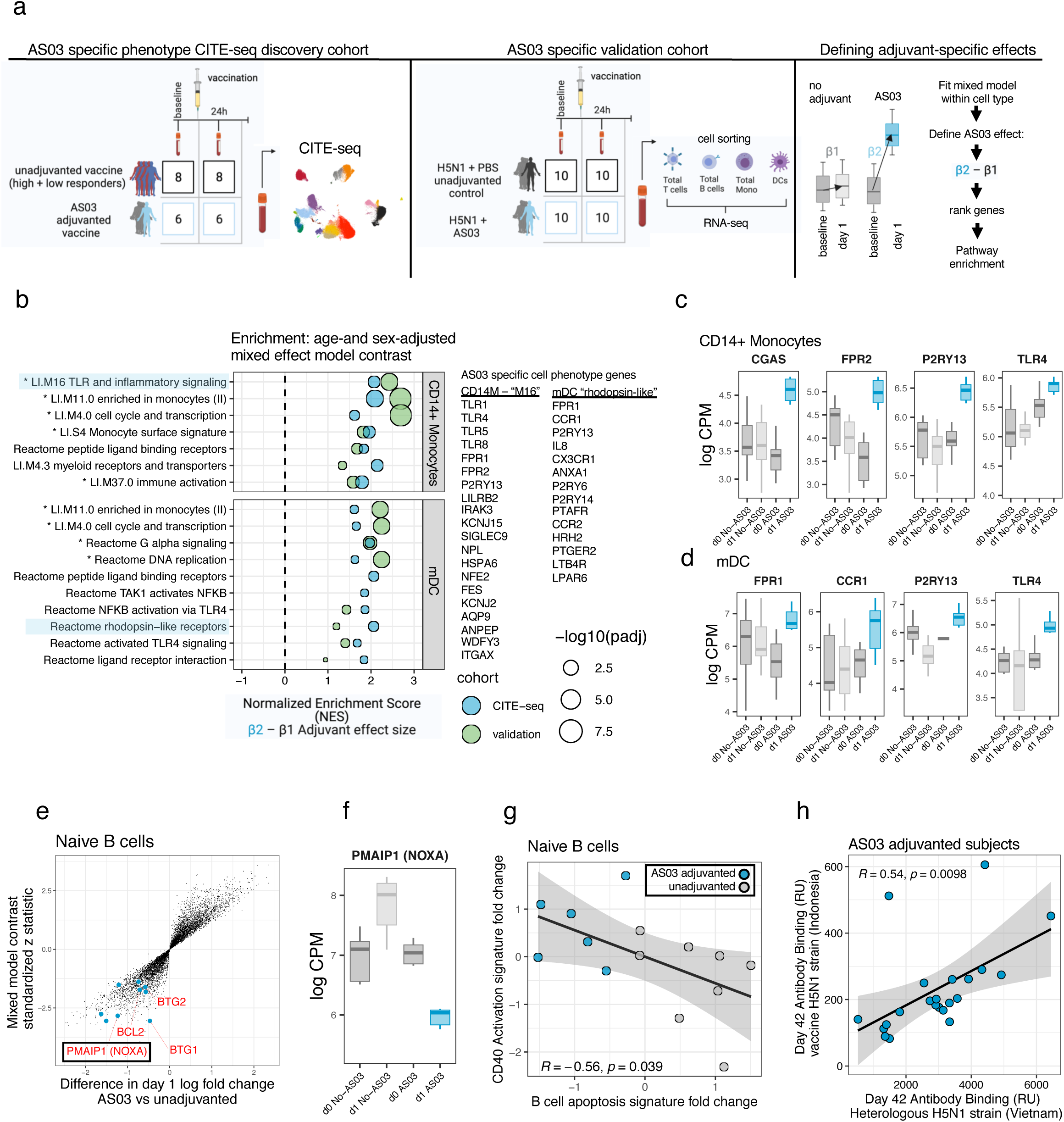
Early transcriptional responses to AS03 adjuvanted vs non-adjuvanted vaccines. **a.** Schematic illustrating the approach to define AS03 adjuvant specific perturbation transcriptome phenotypes within protein based cell types. Left: unadjuvanted individuals were combined and compared to individuals receiving the AS03 adjuvanted vaccine; protein based cell types are the same as those used in Fig 2 and Supplementary Fig 2 which were defined together with the adjuvanted subjects here in combined clustering. Middle: a cohort of individuals vaccinated with AS03 vs an unadjuvanted formulation from Howard *et al.* 2017. Cell types including total T cells, B cells, monocytes and DCs were defined using surface protein and sorted using FACS followed by RNAseq at baseline and day 1. Right: the model contrasts within each cell type applied to the CITE-seq discovery and FACS validation cohorts–for each cell type genes are fit with a mixed effects model and the difference in day 1 fold change between AS03 adjuvanted and unadjuvanted subjects is calculated as shown with boxplots. Genes are then ranked for enrichment based on the effect size of this contrast reflecting AS03 specificity, e.g. modules with positive normalized enrichment score have higher day 1 fold change in the AS03 vaccine group compared to the unadjuvanted vaccine. **b.** Gene set enrichment analysis of genes ranked based on the difference in transcriptional response 24-hours post vaccination vs baseline between AS03+H5N1 vs. H1N1 non adjuvated vaccine (i.e. ranked by the contrast effect shown as in a) in classical monocytes and mDCs. Leading edge genes driving the enrichments of the selected pathways highlighted in light blue are shown to the right. Pathways with adjusted p < 0.01 in the validation cohort are highlighted with an asterisk. **c.** The distribution of log counts per million from aggregated CITE-seq data for each subject of select genes driving difference in perturbation response distinct to AS03 adjuvant within CD14 monocytes. Individual gene statistics from the mixed effects model contrast: FPR2 standardized z: 2.57 p value 0.010, P2RY13 standardized z: 2.56 p value 0.010, MB21D1 (CGAS) standardized z: 2.26 p value 0.022, TLR4 standardized z 1.99 p value 0.047 and **d.** As in c for in mDCs Individual gene statistics from mixed model contrast: FPR1 standardized z 2.91 p value 0.004 P2RY13 standardized z 2.8 p value 0.0051 CCR1 standardized z 2.33 p value 0.02 TLR4 standardized z 1.85 p value 0.0642. **e.** For naïve B cells, the distribution of genes from the mixed effects model showing x axis: estimated difference in baseline vs day 1 log fold changes between AS03 adjuvanted and unadjuvanted vaccination and y axis: standardized z statistics of the fold change difference contrast. Leading edge genes from M160 are highlighted in blue, with additional canonical apoptosis genes not in M160 PMAIP1 (NOXA) and BTG1 highlighted, each with strong AS03-specific downregulation (NOXA standardized z: -2.83, p value: 0.005, BTG1 standardized z: -3.05, p value: 0.002) **f.** Expression distribution of PMAIP1 (NOXA) log counts per million of aggregated CITE-seq data across donors within naïve B cells pre and post vaccination. **g.** Pearson correlation between the day 1 fold change in the CD40 activation score and the apoptosis signature in naive B cells. **h.** The correlation between antibody avidity to the heterologous strain (x-axis – H5N1 Vietnam HA) vs the vaccine strain (y-axis – Indonesia H5N1 HA) (Pearson correlation) measured by surface plasmon resonance assay on day 42 post vaccination in subjects receiving AS03 adjuvant.

While our observations thus far are consistent with the expectation that myeloid cells are key players mediating the effects of AS03, we also detected a lymphocyte signature suggestive of apoptosis suppression in naïve B cells in subjects vaccinated with AS03; this signature included AS03-specific downregulation of genes related to apoptosis (**Figure 3e**). Further examination of genes with the largest difference in post vaccination effects in naïve B cells revealed additional AS03 specific downregulation of canonical pro-apoptotic genes, including BTG1 and NOXA (PMAIP1) (**Figure 3e,f**). NOXA deficiency is known to increase lymphocyte repertoire diversity^64,65^. B cells from NOXA^-/-^ mice outcompete wild type cells for entry into the germinal center following influenza vaccination and infection, and they persist longer due to inefficient apoptosis^65^ and thus increase the diversity of anti-influenza antibodies. As we and others have shown, AS03 induces antibody production against influenza clades beyond those in the vaccine^33,34^. The naïve B cells in humans after vaccination with AS03 may thus phenocopy those in NOXA^-/-^ mice after influenza vaccination. Naïve B cells from subjects vaccinated with AS03 also appeared more activated based on increased expression of genes linked to CD40 activation^66,67^ **(Figure S3c).** The fold-change in the CD40 activation signature score (day 1/day 0) was also negatively correlated with that of an apoptosis signature score in naïve B cells across individuals (**Figure 3g**). Both the apoptosis and CD40 activation signatures had consistent directions of change in sorted total B cells in the validation cohort **(Figure S3d)**, although the apoptosis signature itself was not significant. Together, these observations suggest that AS03 may function to suppress apoptosis in naïve B lymphocytes early after vaccination to prolong their survival and subsequent activation. This potential increase in the diversity of the naïve B cell pool (presumably with varying specificity to vaccine antigens) may help increase the diversity of the subsequent B cell response. We further found that the day 42 antibody avidity to both the vaccine and non-vaccine influenza strains was tightly correlated across individuals immunized with the ASO3 adjuvant (**Figure 3h**), supporting the hypothesis that AS03 may tune the size of the initial naïve B cell pool available to be proportionally expanded in the germinal center. Together these results highlight two potential mechanisms by which AS03 may drive more robust antibody responses: 1) activation of broad innate sensing pathways unrestricted to only those specific to the molecular patterns in the vaccine; 2) suppression of apoptosis in naïve B cells to increase the diversity of naïve B cells entering germinal center reaction with potential positive impacts on antibody response breadth. Detailed information on these AS03 specific cell perturbation phenotypes are provided in **Supplementary Table 2**.

### Linking baseline set point signatures to early vaccination responses reveals natural adjuvanted baseline immune states in healthy humans

We previously described a baseline immune set point signatures predictive of antibody responses to vaccination in healthy individuals and plasma cell-associated disease activities in SLE patients^22^. However, we had only focused on a single class of signatures that was discovered earlier via flow cytometry and bulk transcriptomic analyses; we also did not assess how baseline immune status overlaps with transcriptional and cellular responses early after vaccination. Here we used multivariate models to first perform an unbiased analysis of baseline immune cell phenotypes associated with antibody responses. To understand how these baseline cell phenotypes associated with the high responders were related to one another, we used correlation network analysis. We then further investigated how these baseline phenotypes were linked to early innate responses and how they were correlated with later cellular responses (see Methods). Our first analysis revealed that effector lymphocyte and innate cell phenotypes comprising the baseline predictive signatures could be grouped into several functional categories based on their correlation across individuals. Together these defined a multicellular set point network **(Figure 4a, Figure S4a)**. Interestingly, the phenotypes with the highest “hub”-like properties tended to reflect innate cell surface receptor pathways in CD14+ monocytes and ISG pathways in CD16+ non-classical monocytes **(Figure S4b)**. Full details on the cell phenotypes and genes driving the high responder network phenotype are provided in **Supplementary Table 2**. Two example cell phenotypes from the network are highlighted **(Figure 4b, c)**. Within CD14 monocytes, the “FC receptors and phagocytosis” genes include those encoding Fc receptors (e.g. FCGR3A, FCGR1A, FCGR2A), regulators of cytoskeletal reorganization active during phagocytosis (e.g. PAK1, ARPC5, CFL1, ARF6), and second messenger signaling molecules (PIP5K1A, PIK3CD, AKT1, MAPK12, ARPC2). Remarkably, this monocyte signature was correlated with 27 cell phenotypes elevated in high responders (adjusted p < 0.05) (**Figure 4b**), including both antigen presentation genes in naïve B cells and interferon response genes in CD16 monocytes **(Figure. 4b, bottom).** ISG expression was elevated in a variety of cell types beyond CD16 monocytes, including CD161+ MAIT-like CD8+ T cells **(Figure 4c, bottom)**, within which the level of IFITM1, IFITM2, ISG15 and IFI6 was increased in high responders. These baseline phenotypes were also correlated to the day 7 plasmablast signature score in blood (**Figure. 4d**), which was predictive of antibody responses. Thus, these correlated transcriptional phenotypes at baseline, both within and across cell types, are associated with the extent of day 7 plasmablast and subsequent antibody increases following vaccination.

**Figure 4.**
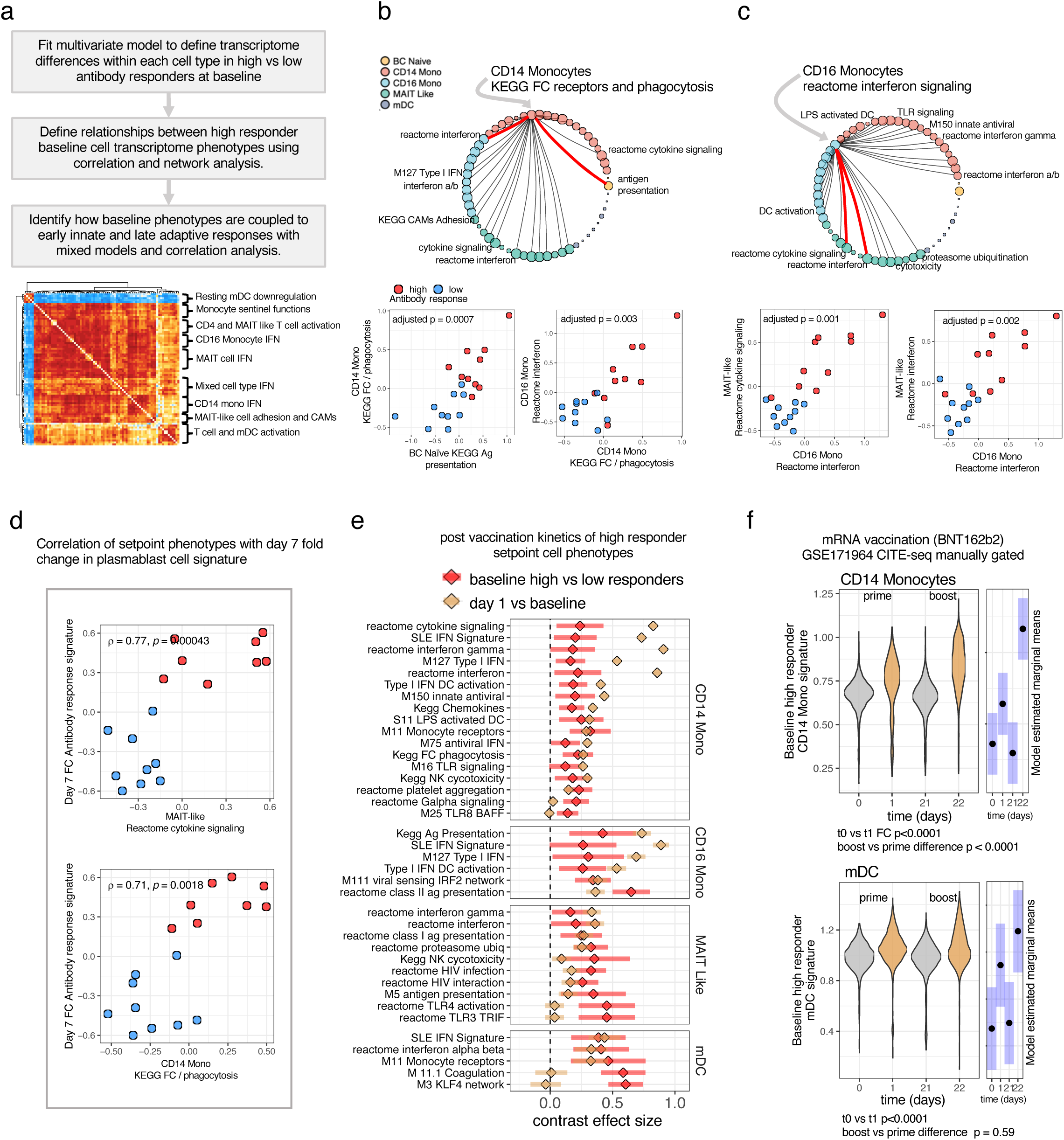
The multicellular setpoint network of high responders their day 1 post-vaccination kinetics and coupling to day 7 plasmablast activity. **a.** Identification of the multicellular baseline high responder setpoint network. Gene set enrichment of modules enriched pre-vaccination (baseline) in high vs. low responders within each cell type based on genes ranked using multivariate models adjusting for age, sex, and batch. The leading edge genes from these cell type specific high responder pathway enrichments were correlated across donors within and between cell types. Within cell types, the Jaccard similarity of each pairwise leading edge gene was subtracted from the spearman correlation coefficient to correct for correlation due to two signals sharing the same genes (within a cell type) and connectivity edges were retained in the network (see methods). **b-c** Two selected highly coupled cell phenotypes in the high responder setpoint network. The edges highlighted in red are shown below as correlations of the activity of the leading edge genes from those modules across donors within the cell type indicated by the edge. Correlation values reflect Bonferroni adjusted Spearman correlation of phenotypes across the entire network. **d.** The correlation of signature expression within cell types with the day 7 fold change in the predictive signature we previously found was predictive of antibody response associated with plasmablast activity from microarray data. **e.** The post vaccination kinetics of the components of the high responder innate setpoint network. A single cell mixed effects model of module activity was used to estimate the baseline high vs low responder effect size (red) and day 1 fold change across subjects adjusting for age, sex, number of cells per donor and a random effect for donor ID. **f.** Day 1 vs 0 prime and day 22 vs 21 boost kinetics of baseline high responder states tested in an external cohort of monocytes and DCs manually gated from CITE-seq data (GSE171964) collected on individuals vaccinated with mRNA vaccine BNT162b2. The difference in the fold change between boost (d22 vs d21) and prime (d1 vs d0) p values: mDC 0.59, CD14 monocyte < 0.001 and day 1 vs baseline p<0.001 calculated by the emmeans package based on a mixed model with a donor random effect as in e.

Interestingly, the phenotypes comprising the above baseline set point network and the innate signatures induced early following unadjuvanted (i.e., without AS03) vaccination (see **Figure 2a**) appeared similar. We thus asked whether the cell phenotypes comprising the high responder set point network were induced by vaccination by statistically modeling the early (day 1) post–vaccination response of the baseline set point signature genes in a cell type specific manner. This analysis revealed the same phenotypes driving this multicellular high responder set point (including CD14 and CD16 monocytes, mDCs, and MAIT cells), were induced by vaccination coherently across all individuals within the same cell subsets (**Figure 4e**).This suggests that the high responder baseline set point signature indeed reflected an immune state mirroring the early inflammatory responses induced by vaccination. This baseline state may have primed innate responses to vaccination since it was itself further induced by vaccination. Further supporting this idea, the baseline signature in monocytes and mDCs was also induced one day after either dose of BNT162b2 mRNA SARS-Cov2 vaccination^68^, with greater elevation after the second dose in classical monocytes (**Figure 4f**). Given that the lipid nanoparticle carrier in the mRNA vaccine is thought to act as an adjuvant^69^, these results further suggest that the baseline set point signatures might have reflected a naturally “adjuvanted” state that can enhance innate immune response potential prior to stimulation.

Interestingly, we noticed that the aggregated AS03 specific early response phenotypes (the union of leading edge genes driving the gene set enrichments in each cell type shown in Figure 3b) were decreased rather than increased after unadjuvanted vaccination, further demonstrating that they were unique to the response to AS03 (**Figure 5a,b**). To further test the naturally adjuvanted baseline hypothesis, we next tested whether these specific DC and monocyte signatures specifically induced by the AS03 adjuvant were phenocopied by the baseline of high responders. Indeed, these AS03-specific innate response phenotypes were higher at the baseline of high than low responders to the unadjuvanted vaccine (**Figure 5c**). A previous study of AS03 identified increased frequencies of activated HLA-DR+ monocytes 24h following vaccination^9^. Again, here the high responders to unadjuvanted vaccination already had elevated frequencies of HLA-DR+ monocytes^6^ at baseline (**Figure 5d**). Furthermore, by day 1 post vaccination, the frequency of these activated HLA-DR+ monocyte increased with a larger effect in the high responders (effect size 3.17, p = 0.0005) than the low responders (1.89, p = 0.14) (**Figure 5e**). Thus, multiple lines of evidence, including those from transcriptional and innate immune cell frequency analysis, support the conclusion that the baseline immune statuses of high responders correspond to a naturally adjuvanted innate immune state that mirrors not only the early responses induced by the unadjuvanted vaccine, but also those specifically elicited by the AS03 adjuvant.

**Figure 5.**
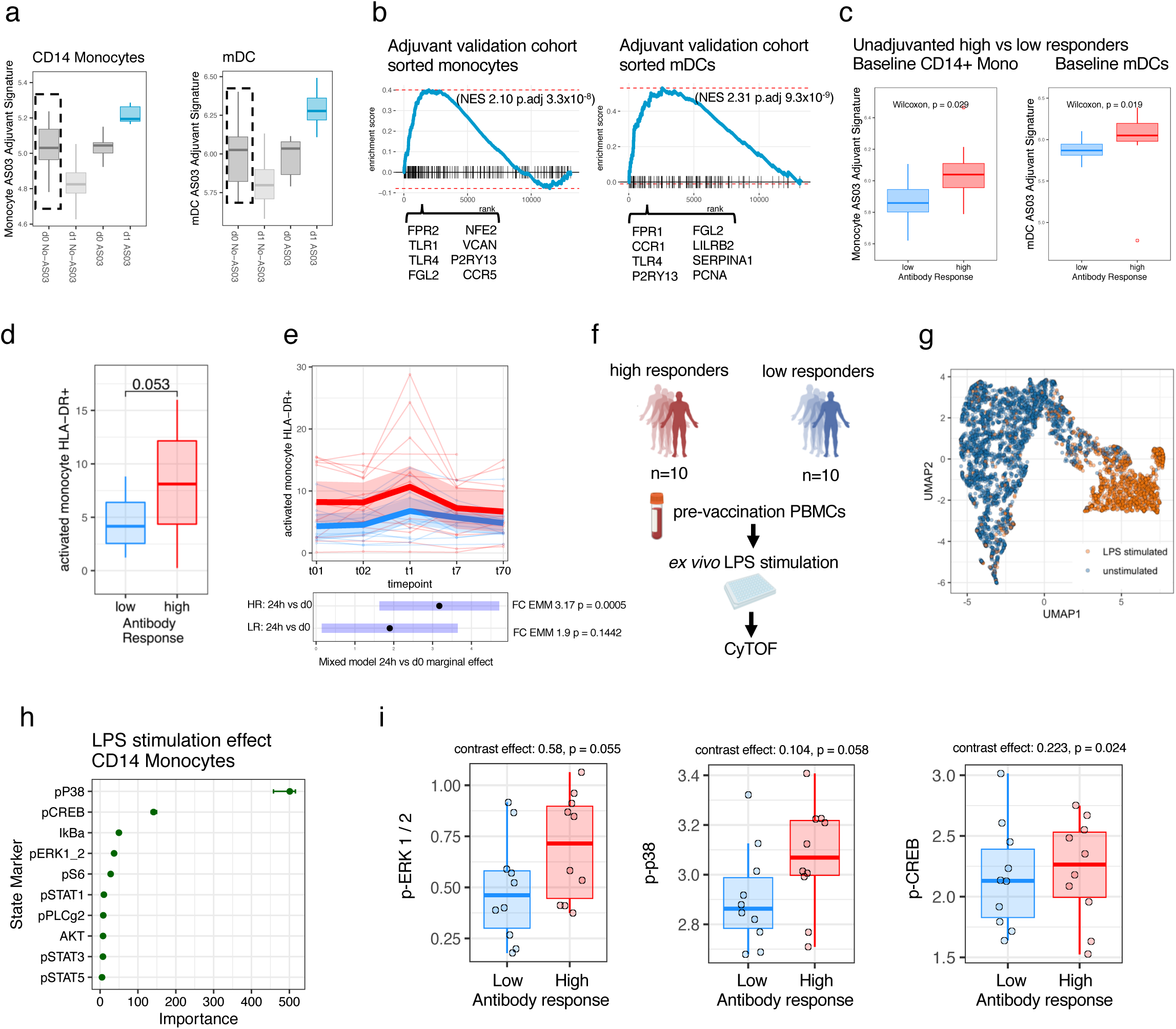
High responders have a naturally adjuvanted immune setpoint with monocytes more poised to enter blood and respond to PRR stimulation. **a.** Average expression of a combined gene signature reflecting the AS03 specific induced states within DCs and CD14 monocytes. **b.** Gene set enrichment of the combined AS03 specific signature on the validation cohort in analogous subsets; select adjuvant specific genes in the leading edge of the validation are shown. **c.** The average expression in high vs low responders of the mDC and CD14 monocyte AS03 specific day 1 induced validated signature tested in analogous subsets. **d.** Log cell frequency of HLA-DR+ classical monocytes as a percentage of total classical monocytes in high vs low responders at baseline, p value from a Wilcoxon rank test. **e.** The kinetics over two baseline timepoints and three post vaccination timepoints for HLA-DR+ classical monocytes. Mixed effects model with an interaction for time and response group and a random effect for subject ID–high responder effect size 3.17 p value = 0.0005, low responder effect size 1.89, p value = 0.14, difference in estimated marginal day 1 vs baseline fold change not significant, response time vs time only interaction model ANOVA p = 0.063. **f.** Schematic outlining CyTOF stimulation experiment. PBMCs isolated from high and low responders were stimulated with PRR ligands. Stimulation phenotype and markers driving stimulation were defined with HDStIM. **g.** UMAP plot of a random subset of 5000 monocytes pre and post stimulation with stimulated cells in orange and unstimulated cells in blue. **h.** Variable importance for individual phospho-protein markers determined by the Boruta algorithm which are used for automatic determination of responding cells in HDStIM. **i.** The post stimulation median marker intensity of phosphor markers within the CD14 monocyte cluster, the post stimulation aggregated data are shown due to variable baseline phospho-marker detection and effects were tested using a mixed model adjusting for batch and modeling individual variation with a random effect for donor ID. The difference in pre vs post stimulation fold changes in high vs low responders contrast estimate and p values: p38 contrast effect: 0.104, p = 0.058, pCREB contrast effect: 0.223, p = 0.024, pERK contrast effect: 0.58, p = 0.055.

The naturally adjuvanted baseline statuses may partly reflect cell-intrinsic differences in response capacity to innate immune cell stimulation. To evaluate this hypothesis, we stimulated PBMCs from the same 10 high and 10 low responders (to the unadjuvanted influenza vaccine) with interferon alpha, PMA plus ionomycin, and LPS, and used early phosphorylation signaling responses within 15 minutes after stimulation to assess whether certain cell subsets were intrinsically more responsive in transducing these external stimulatory signals (**Figure 5f**). We used CyTOF profiling for both cell surface protein and intracellular phosphorylation-based signaling readouts, and defined the responding cell populations and associated response markers by using a computational algorithm we recently developed called HDStIM^70^ (**Figure 5g-h**). As expected, CD14 monocytes responded strongly to LPS as evident by increased levels of phosphorylated p38, CREB, IkBa, and ERK (**Figure 5h**). Supporting the idea that the naturally adjuvanted set point reflected cell intrinsic signaling response capacity, the difference in the post-stimulation fold-change of p38, pERK, and pCREB (after adjusting for batch and individual variation) was elevated to a greater extent in high compared to low responders (**Figure 5i**). This cell intrinsic, TLR-dependent increase in the signaling capacity of monocytes suggest that the high responders possess a baseline set point poised to mount a stronger response to stimulatory signals from the vaccine. Specifically, if this intrinsic signaling response difference extends to pattern-recognition receptors that might recognize influenza vaccine components, such as TLR3, TLR7, or TLR9, these may signal through transcription factors including IRF3 or IRF7 to activate interferon response genes, such as those encoding for ISG15 and IFN-β; these could further induce antiviral gene expression programs in both monocytes and DCs via autocrine / paracrine circuits^71,72^. Furthermore, enhanced p38 signaling could also play a role in RIG-I induced interferon response to the vaccine^73,74^.Together these observations provide additional insights into the mechanistic underpinnings of a naturally adjuvanted human immune set point found in healthy individuals primed to respond with more robust innate and adaptive responses following vaccination.

## Discussion

In this work we introduce a framework for integrating natural human population variation with multimodal single cell variation capturing cellular states before and after a perturbation. While prevailing analysis approaches for single cell data often rely on qualitative visualization^75^ and univariate analysis, these approaches are often insufficient for complex experiment designs with many samples^76^ and do not provide a quantitative means to integrate human and single cell variations to extract biological insights. Our approach provides robust statistical methods for these complex experimental designs; its application to the multimodal single cell data in this work illustrated how new insights can be obtained, e.g., regarding adjuvant specific response phenotypes involving naïve B cells as well as the cellular and transcriptional signatures of a naturally-adjuvanted baseline immune set point. These findings advance the concept that modulating baseline set points may improve immune response outcomes in diverse contexts^21^. For example, the baseline immune states of the low responders could be tuned to phenocopy the naturally adjuvanted innate immune state to enhance their future vaccination responses; these low responders include patients who require continued immunosuppression e.g. after transplantation, but need urgent vaccination in a pandemic setting.

A host of approaches can be used to tune immune set points including vaccination itself. For example, BCG vaccination has been known to confer nonspecific protection (i.e., not just against TB) and reduce all-cause mortality in infants^77^; it has also been shown to potentiate nonspecific secondary innate immune cell responses in mice^78^. Recent phase III human trials evaluating BCG vaccination as a nonspecific immunomodulator showed promise in demonstrating protection against respiratory infections in the elderly^79^, who tend to be immunosuppressed^43^. It remains to be seen whether the naturally adjuvanted phenotype we describe here is similar to the innate immune training conferred by BCG vaccination^80,81^, which can induce short-term innate immune memory attributed to chromatin remodeling^82^. Indeed, the molecular underpinnings of the naturally adjuvanted baseline transcriptional phenotype remain to be determined. Preliminarily by using a computational approach^83^ to look for transcriptional factor motif enrichments, we detected significant enrichment of SPI1/ PU.1, IRF family members, and CEBPB, which were ranked near the top among other transcription factors (TFs) predicted to regulate the above set point signature genes in classical monocytes (these were ranked between 2 and 32 among 1632 tested, data not shown). Intriguingly, these were some of the same TFs whose binding motifs tended to have altered chromatin accessibility after LPS “training” in mouse monocytes, leading to enhanced myelopoiesis and elevated extravasation of monocytes into the blood^84^. Future work could evaluate vaccination regimens which might optimize the longitudinal persistence of this naturally adjuvanted set point.

Evaluation of larger cohorts using similar multimodal single cell approaches will help assess the generalizability of our naturally occurring baseline set points. While lacking the resolution of the multimodal single cell analysis framework introduced here, our earlier work analyzing bulk blood transcriptome data from multiple influenza vaccine studies provide independent support, including the observation of substantial inter-subject variation in baseline immune states^1,2,6,85^ and an “inflammatory signaling” module predictive of antibody response to influenza vaccination in multiple cohorts of subjects under the age of 65^16^. More recent work using bulk blood transcriptomic data assessing different types of vaccines revealed that individuals with a high “inflammation” phenotype tended to have better antibody responses^23^. What is less clear is how age related inflammation is similar to or distinct from such baseline inflammatory states. Earlier work suggests that tonic levels of interferon in the young are distinct from age related inflammation, which may be more related to TNF signaling and its downstream effects^86,87^. Our work provides a basis for future studies to identify the extent by which these bulk signatures can be resolved further by using the kind of approaches introduced here.

Our study has several limitations. Profiling blood alone misses cells and processes in tissues. Assessing tissues such as lymph nodes would give a more comprehensive picture of vaccination response variations across individuals. Despite logistical challenges of human tissue profiling, recent pioneering work using fine needle aspirates or biopsies from lymph nodes^88–90^ following influenza vaccination^91^ have helped link blood and tissue phenotypes. For example, our single cell deconvolution revealed that the predictive day-7 bulk expression signatures were derived nearly exclusively from a small number of plasmablast cells (Figs S2c-e). Circulating plasmablasts have been shown to shared B-cell receptor sequences with those obtained from lymph node biopsies^91^, thus the whole blood based plasmablast transcriptional signatures that have been widely detected post vaccination in previous studies are, as expected and supported by our results, originated from B cells in lymph nodes with shared clonality. Determining the origin of the innate immune cells and their states in circulation, including both DCs and monocytes, on day 1 and their connection to the cells “encoding” the naturally adjuvanted baseline states remains an open problem. Given that monocytes have relatively short halflife, the dynamics and status of the myeloid progenitors need to be considered and may hold a key to linking immune cell status in the bone marrow and shorter-lived circulating cells in blood. Tracking the clonal origins of innate immune cells lacking clonal receptors in humans presents a major challenge, however, recent developments in mitochondrial DNA mutation profiling using single cell ATAC-seq data could be informative in this context^92^. Another open issue is the origin of the naturally adjuvanted baseline immune state within individuals – what sets the set point? Our recent work suggest that prior infections could modulate and establish new baseline set points in humans, e.g., months after clinical recovery from mild COVID-19 both men and women had a temporally stable altered baseline immune state compared to matching controls, and men tended to mount more robust innate and adaptive responses to the seasonal influenza vaccine^93^. As future work we can assess whether and how the monocyte and DC naturally adjuvanted phenotypes overlap with those stably modified by prior infections in the same cells. Finally, vaccination itself, such as BCG discussed above as well as recent evidence from influenza vaccination with adjuvants^94^, can also potentially modulate baseline immune states. Together, our framework paves the way for further studies to integrate human and single cell variations over space and time in response to perturbations across biological disciplines; our findings help advance a more quantitative, predictive understanding of the human immune system.

## Methods

### Human vaccination comparison cohorts and antibody response assessment

Healthy volunteers were enrolled on the National Institutes of Health (NIH) protocols 09-H-0239 (Clinicaltrials.gov: NCT01191853) and 12-H-0103 (www.clinicaltrials.gov: NCT01578317). Subjects enrolled in 09-H-0239 received the 2009 seasonal influenza vaccine (Novartis), and the 2009 H1N1 pandemic (Sanofi-Aventis) vaccines, both without an adjuvant. Subjects in 12-H-0103 received a vaccine formulated with the adjuvant AS03 containing avian influenza strain H5N1 A/Indonesia/05/2005 (GSK). In both cohorts, virus neutralizing antibody titers assessed using a microneutralization assay were determined as previously reported. The highest titer that suppressed virus replication was determined for each strain in the 2009 inactivated influenza vaccine: A/California/07/2009 [H1N1pdm09], H1N1 A/Brisbane/59/07, H3N2 A/Uruguay/716/07, and B/Brisbane/60/2001 or for AS03 adjuvanted influenza vaccine, H5N1 A/Indonesia, clade 2.1. High and low antibody responders to the unadjuvanted vaccination were defined using the adjusted maximum fold change (AdjMFC) which adjusts the fold change for the baseline antibody titer (methodological details in the supplementary methods of our previous report^6^). In the unadjuvanted cohort, n=10 high responders and n=10 low responders were selected for CITE-seq profiling. All subjects were analyzed pre–vaccination, with a subset of 8 and 12 donors profiled on days 1 and 7 post-vaccination also split evenly between high and low responders. In the adjuvant cohort, n=6 subjects with robust titer responses were selected for CITE-seq.

### CITE-seq profiling of peripheral blood mononuclear cells

We optimized a custom CITE-seq antibody panel of 87 markers using titration experiments and stained cells with a concentration of antibody appeared to saturate ligand of the cell population with the highest marker expression, or used the manufacturers recommended concentration when below saturation. We stained the 52 PBMC samples across three experimental batches using a single pool of which were combined in the optimal concentration and concentrated in an Amicon Ultra 0.5mL centrifugal filter by spinning at 14,000 x g for 5 minutes. Three aliquots of 12µL from the 36µL volume of optimized antibody mixture was used on 3 subsequent days to minimize between experiment technical variability. Frozen PBMC vials from each donor were washed in pre-warmed RPMI with 10% FBS followed by PBS. 1x10^6^ cells from each sample were stained with a hashing antibody^95^ simultaneously with 1µL FC receptor blocking reagent for 10 minutes on ice. After washing the hashing reaction 3 times in cold PBS, cells were counted and pooled in equal ratios into a single tube and mixed.

The sample pool was concentrated to 5x10^6^ cells in 88µL of staining buffer. 12µL of the concentrated optimized 87 antibody panel was added to stain cells (total reaction volume 100µL) for 30 mins on ice. After washing cells, we diluted cells to 1400 cells / µL, recounted 4 aliquots of cells and 30µL of the stained barcoded cell pool containing cells from all donors was partitioned across 6 lanes of the 10X Genomics Chromium Controller for each of the 3 batches for 18 total lanes. We proceeded with library prep for the 10X Genomics Chromium V2 chemistry according to the manufacturer’s specifications with additional steps to recover ADT and HTO libraries during SPRI bead purification as outlined in the publicly available CITE-seq protocol (https://cite-seq.com) version 2018-02-12. We clustered Illumina HiSeq 2500 flow cells with V4 reagents with pooled RNA, ADT and HTO libraries in a 40:9:1 ratio (20µL RNA, 4.5 µL ADT, 0.5µL HTO). Libraries were sequenced using the Illumina HiSeq 2500 with v4 reagents. CITE-seq antibody information is provided in Supplemental Table 5.

### CITE-seq data sequence alignment and sample demultiplexing

Bcl2fastq version 2.20 (Illumina) was used to demultiplex sequencing data. Cell Ranger version 3.0.1 (10x Genomics) was used for alignment (using the Hg19 annotation file provided by 10x Genomics) and counting UMIs. The fraction of reads mapped to the genome was above 90% for all lanes and sequencing saturation was typically around 90%. ADT and HTO alignment and UMI counting was done using CITE-seq-Count version 1.4.2. We retained the “raw” output file from Cell Ranger containing all possible 10X cell barcodes for each 10X lane, and merged the CITE-seq-count output. For each 10X lane, barcodes were concatenated with a string denoting the lane of origin and data for ADT, HTO and mRNA. We then utilized combined sample demultiplexing to assign the donor ID and timepoint to each single cell. Both the timepoint and response class were identifiable based on the hashing antibody. The first round of demultiplexing was carried out via cell hashing antibodies. The union of singlets defined by the multiseq deMUTIplex procedure^96^ and Seurat’s HTODemux function were retained for further QC. Negative drops identified by HTODemux were retained for further QC and use in denoising and normalizing protein data. The second round of sample demultiplexing was carried out via Demuxlet^97^ to assign the unique donor ID by cross-referencing unique SNPs detected in mRNA single cell data against a vcf file with non-imputed illumina chip based genotype data from the same donors. Demuxlet provided an additional round of doublet removal via an orthogonal assay (mRNA) to antibody barcode (HTO) based demultiplexing thus providing further data QC. Only cells that met the following conditions were retained for further downstream QC, normalization and analysis: 1) The cell must be defined as a “singlet” by antibody barcode based demultiplexing and by demuxlet. 2) The identified donor from demuxlet must match one of the expected donors based on cell hashing. Cells were then further QCd based on mRNA using calculateQCmetrics function in scater^98^. Cells were removed that had with greater or less than 3.5 median absolute deviations from the median log mRNA library size.

### Surface protein and mRNA count data normalization

We denoised and normalized ADT data using an open source R package we developed for this work called dsb^37^ which removes noise derived from ambient unbound antibodies and cell to cell technical noise. We used function DSBNormalizeProtein with default parameters. We normalized mRNA on the entire dataset with the normalizeSCE and multiBatchNorm functions from scran^99^ using library size-based size factors. Various analysis utilized aggregated mRNA data which was were separately normalized for analysis at the subset level as a “pseudobulk” library; single cell mRNA data were also renormalized or rescaled for specific analysis as outlined below.

### Surface protein-based clustering and cell type annotation

Using protein to define cell type facilitated improved interpretation of transcriptome differences between vaccination groups. Cell types were defined with statistically independent information, protein, from transcriptome data being modeled within each cell type (Figure. 1a). We clustered cells directly on a distance matrix using the parallelDist package calculated from the non-isotype-control proteins all cells using Seurat’s FindClusters function using parameters: res =1.2, modularity.fxn = 1, algorithm = 3 (SLM^100^). We annotated cell types in the resulting clusters post hoc, based canonical protein expression in immune cell populations. This procedure improved separation of known immune populations compared to compressing protein data using principal components as commonly done for higher dimensional mRNA data (data not shown). Analysis of unadjuvanted vaccination responses was first done blind to the adjuvanted cohort data. We thus first applied high dimensional clustering of the unadjuvanted cohort and annotated cell types with additional manual gates to purify canonical cell populations such as memory and naïve T cells. We next merged unadjuvanted and adjuvanted cohort cells and used annotations to guide combined clustering annotation, again manually refining cell populations using biaxial gating scripts in R to purify cell some cell populations. For annotation, the distribution of marker expression within and between clusters was compared using density histogram distributions of marker expression across clusters at the single cell level, biaxial marker distribution and median and mean aggregated protein expression across clusters.

### Hierarchical transcriptome variance deconstruction to infer individual (subject intrinsic), cell type, and vaccine effects

To estimate the contribution of subject intrinsic and contributors to the observed variation in expression of each gene within specific cell clusters/subsets, we used the variancePartition package^101^. The set of models used for estimating variance fractions are distinct from but related to those used for testing differential expression and contrast vaccination effects within cell subsets (see below). We first aggregated data across individual, timepoint and cell type. The normalized aggregated expression was used to first model the mean variance relationship using observation level weights using voom^102^. Mixed effects linear models of the expression of each gene across the aggregated libraries were then fitted using lme4^103^ with variancePartition. For each gene “y” the total variance was defined by 780 measurements derived from the 52 PBMC samples deconvolved into the 15 major protein-based cell clusters/types tested. The model fit to each gene “g” was:

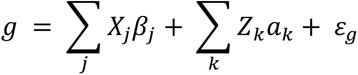

Where X and Z are the matrices of fixed and varying / random effects respectively, with random effects modeled with a Gaussian distribution and errors incorporating weights calculated with voom.

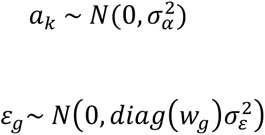

The variancePartition package then incorporates both fixed and random effects in calculating the fraction of variation attributable to each variable in the model. For example, the variance in g attributable to “subjectID” (i.e., differences between individuals) was modeled as a random effect is:

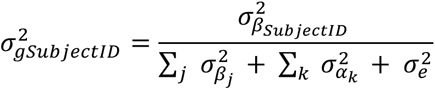

The denominator in the fraction above is the total variance of gene g, with both fixed and random effects contributing to total variance. In the first model above, age, sex, subjectID, timepoint, response /vaccine group (unadjuvanted group high vs low responders, or AS03 group) cell type, and a cell type and timepoint interaction term as categorical random effect variables as required by the variancePartition framework. As expected, a second set of models fit within each cell type/cluster (i.e., without having cell type as a variable in the model) increased the apparent variance explained by the other factors given that major cell type specific expression was a key factor driving gene expression variation. This model included age, sex, subjectID, timepoint, and response / vaccine group (as above) and an interaction term for time and group.

### Within cell type linear mixed effect models of vaccination effects on gene expression

We used linear mixed models to test coherent effects of vaccination across individuals while adjusting for subject intrinsic factors including age and gender and estimating individual subject level variation. Gene expression counts were aggregated within each surface protein-based cell type by summing counts within each sample. The lowest frequency cell types without representation across some individuals and time relative to vaccination (e.g., HSCs, donor-specific cell types, or plasmablasts which were mainly detected on day 7) were excluded from this specific analysis. Three main analysis were carried out to model gene expression within each cell type to estimate the following vaccination effects over time across individuals: model 1) unadjuvanted subjects day 1 vs baseline, model 2) unadjuvanted subjects day 7 vs baseline, model 3) A contrast of the difference in day 1 fold change between unadjuvanted and adjuvanted subjects in a combined model – the goal of this model is to assess adjuvant specific response effects. All models were fit with the ’dream’ method^96^ which incorporates precision weights^97^ in a mixed effects linear model fit using using lme4^98^. For models 1 and 2 above (unadjuvanted vaccination effects) we fit the following model: *gene ∼ 0 + time + age + sex + (1|subjectID)*.

The fitted value for expression y of each gene g corresponds to:

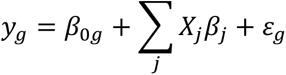

With variables time, age, and sex represented by covariate matrix *X*. The *β*_0_ term corresponds to the varying intercept for each donor represented by the (1|subjectID) term. This model thus estimates the baseline expression variation across subjects *S*_0_ around the average *γ*_0_ using a Gaussian distribution with standard deviation τ_g_^2^ to shrink estimated vaccination effects toward the population mean and adjust for non-independence of repeated measures from the same individuals, as follows:

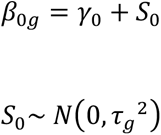

Errors *ε_g_* incorporate observational weights *w_g_* calculated using the function *voomWithDreamWeights* in a procedure similar to that described by Law *et al*^102^ but using the mixed model fit:

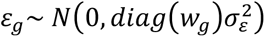

In this model, the day 1 or day 7 effect across subjects was the time effect from the model. The mixed model standardized z statistic was then used to rank genes for gene set enrichment testing for each cell type. Model 3 was specified as *gene ∼ 0 + group + age + sex + (1|subjectID)*. The “group” variable corresponds to a combined factor representing the vaccine formulation received (adjuvanted vs unadjuvanted) and timepoint (baseline or day 1 post vaccination) with 4 level: “d0_AS03”, “d1_AS03”, “d0_unadjuvanted”, “d1_unadjuvanted”. A contrast matrix *L_delta_* corresponding to the difference in fold changes between adjuvanted and unadjuvanted subjects was applied to test the null hypothesis of 0 difference in fold changes between the groups.

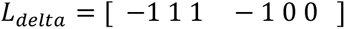

With the first four columns representing the group factor and the two 0s representing age and sex effects. The contrast fit outputs the difference in fold change after adjusting estimates for age, sex and subject variation with positive effects representing increased fold change in the adjuvant group compared to the unadjuvanted group. This contrast approach was designed to also capture genes with opposite vaccination effects in the two groups, for example, upregulation in the AS03 group and downregulation in the nonadjuvanted subjects.

Transcriptome data was uniformly processed for all fitted models above. Aggregated (summed) single cell UMI counts were normalized within each protein based cell type using the trimmed means of M values method with only genes retained with a pooled count per million above 3 using the edgeR *filterByExprs* function^104^. Cell type specific gene filtering removed genes non expressed by each lineage from analysis ensured the model assumptions used to derive precision weights and account for the mean variance trend were met. We verified the log count per million vs. fitted residual square root standard deviation had a monotonically decreasing trend within each cell type. For the AS03 validation cohort, pre normalized data were downloaded from the study supplemental data^58^ and a similar model to model 3, contrasting the difference in fold change was fit with a contrast again using a donor random intercept.

### Gene set enrichment testing of vaccination effects within cell types using specific hypothesis-driven gene sets or unbiased analysis

To test enrichment of pathways based on the estimated gene coefficients corresponding to the three vaccination effects defined above, we performed gene set enrichment analysis using the fgsea^105^ package multilevel split Monte Carlo method (version 1.16.0). Genes for each coefficient (i.e. models 1-3) and each cell type were ranked by their effect size, (the dream package empirical Bayes moderated signed z statistic), corresponding to pre vs post vaccination or the difference in fold change for model 3 (comparing unadjuvanted vs. AS03). For enrichment of the day 1 response, five gene sets were derived from bulk transcriptomic data of influenza vaccination (see Supplementary Table 1), and an additional 25 pathways/gene sets curated from public databases were tested. For Day 7 responses and the difference in fold change between adjuvanted and unadjuvanted subjects, an unbiased set of pathways were tested from the Li et al. Blood Transcriptional Modules (BTM)^106^, MSigDB Hallmark, reactome and kegg databases. Over-representation of GO terms for the monocyte pseudotime gene categories was assessed using enrichr^107^.

### Inference of the baseline immune set point network

To define cell type specific transcriptional phenotypes robustly associated with high vs low responders of the unadjuvanted vaccine at baseline, we used limma^108^ to fit linear models of gene expression as a function of antibody response class (high vs low, coded as a two-level factor) adjusting for age sex and batch (e.g. in R symbolic notation, gene ∼ AdjMFC + age + sex + batch) as fixed effects on aggregated (summed) data for each cell type, similar to models above without varying effects for individuals:

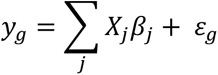

Errors incorporated voom weights as above. Gene coefficients for each cell type corresponding to model adjusted empirical Bayes regularized estimates for high vs low responder effect at baseline were input into gene set enrichment analysis against the unbiased set of pathways described above. We then calculated the average module z score^22^ using log counts per million from each cell type of the high responder associated cell phenotypes (using only high responder associated leading edge genes from gene set enrichment analysis), resulting in a matrix of baseline normalized expression of pathways across 20 individuals (10 high and low responders) for each cell type. We next tested for correlation of these signals, both within and between cell types, by calculating the spearman correlation and adjusted p values with the FDR method. We noticed that within the same cell type, pathway enrichments could sometimes be driven by a shared set of genes among gene sets with different pathway labels but essentially shared a substantial fraction of genes. We therefore calculated the Jaccard similarity coefficient of each pairwise enrichment signal (leading edge genes driving the high vs low responder difference) within each cell type, and use that to adjust the correlation effect sizes computed above such that the resulting quantity reflected “shared latent information” (SLI) by subtracting the Jaccard similarity index from the Spearman correlation coefficient ρ:

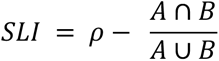

For example, given enriched pathways A and B within a cell type, if at one extreme, these two pathways are driven by the same exact shared 10 leading edge genes, the Spearman ρ of their normalized expression would be equal to 1, yet this apparent correlation is arbitrary since the two pathways reflect the same genes. However, the shared latent information would be equal to 0 because the Jaccard similarity of the two sets is also equal to 1 since the leading edge genes from the enrichments are also the same. The remaining correlation strength better reflects the phenotypic coupling of intracellular states across individuals after removing the signal due to leading-edge gene sharing between gene sets. For inter-cellular correlations between two distinct cell types, we do not subtract the Jaccard similarity of gene content from ρ as we consider the same genes to be distinct signals when measured in different cell types. We further constructed a sub network from a subset of cell types forming the high responder baseline set point network. To identify the most highly connected processes, correlations with adjusted p values < 0.05 were retained and a weighted undirected network was constructed using igraph, retaining only the strongest links above the median weight with weights reflecting Spearman’s Rho for intercellular connections and the SLI metric described above for intracellular connections. Each node (high responder cell phenotype) was also correlated across individuals with the day 7 fold change of a gene expression signature^5^ reflective of plasmablast activity derived from bulk microarray data from the same subjects and select high degree nodes were highlighted in the text.

### Single-cell mixed-effect models of gene expression

In addition to the pseudo-bulk models fitted above, we also used single cell mixed effects models to assess consistency and to specifically test the early response kinetics of the baseline states enriched above, including select AS03 associated response signatures within innate immune cell subsets.

#### Early kinetics of baseline set point phenotypes

Each cell type specific transcriptional phenotype enriched in high vs low responders in the aggregated/pseudo-bulk linear model described above were scored in single cells from subjects on day 0 and day 1 as the average expression of the specific leading edge genes enriched in high vs low responders. The per single cell module scores were fitted with a linear mixed model for each cell type to 1) re-test the baseline association (high vs. low responders) at the single cell level, and 2) to test their post vaccination effect size within the same cell subset. These models estimated the variance at the single-cell level instead of at the individual donor cell-aggregated level. Otherwise these represent conceptually similar models as the ones described above fitted using lme4 with a donor random intercept, but without voom weights. Two models were tested with highly concordant resulting effect sizes: 1) a parsimonious model of time relative to vaccination with a subject random effect, and 2) a more complex model including the time relative to vaccination, the number of cells per individual sample for a given cell type, age, sex, and a subject random effect. Normalized expression of each module was standardized within each surface protein-based cell cluster/subset by subtracting the mean and dividing by the standard deviation of the module score across single cells within the cell type. After fitting models, the baseline high vs low responder effect and the day 1 vs baseline effect sizes and standard errors across subsets was calculated using the emmeans^109^ package with a custom contrast (e.g., see Figure. 4e). All models were checked for convergence criteria.

#### AS03 specific regulation

Naïve B cells were tested for expression of modules hypothesized to be involved in B cell survival (see below; partly based on the literature or derived from existing independent data sets). These modules were tested here for their effects at the single cell level; they were then independently assessed in sorted total B cells in the validation cohort^58^. Two modules were defined to reflect survival of human naïve B cells: 1) A CD40 activation signature^22^ which was derived from studies of in vitro CD40 activated human B cells; 2) An apoptosis signature derived by combining signals from the CITE-seq naïve B cell day 1 gene set enrichment comparing AS03 adjuvanted to unadjuvanted individuals. The signals combined the specific naïve B cell leading edge genes from the negatively enriched (reflecting AS03 specific downregulation) apoptosis modules (with unadjusted p values < 0.1–we opted for a loser cutoff to increase sensitivity): reactome activation of BH3 only proteins, KEGG p35 signaling pathway, and LI.M160 leukocyte differentiation. The cell type specific leading edge genes were scored as above and fitted with age and sex covariates, a combined factor for vaccine group, timepoint, and random effect for subject ID, with the difference in fold changes calculated using the emmeans package.

### Software for implementing analysis workflow

The analysis framework described above is available in an R software package “scglmmr” (https://github.com/MattPM/scglmmr) for analysis of single cell perturbation experiment data with repeated measures and multi-individual nested group designs. The software provides workflows for fitting single cell mixed models, deriving cell signatures, visualization, and also includes wrapper functions to implement the weighted gene level mixed effects differential expression models described Hoffman *et al*. 2021 (dream) and enrichment using fgsea.

### Monocyte differentiation and perturbation pseudotime analysis

To construct a combined monocyte differentiation and perturbation single cell map we used the DDR tree algorithm with monocle 2^57^. The trajectory was constructed using the genes that changed as a function of time (q value <0.15 using the differentialGeneTest in monocle, with ribosomal genes and genes expressed in less than 15 cells removed). The DDRtree algorithm^56^ was implemented using the monocle function reduceDimension with arguments *residualModelFormulaStr* = subjectID and *max_components* = 2 and pseudotime calculated with function orderCells. Independently of the genes used to construct the trajectory, we then tested the genes from the mixed effects model of vaccination effects from monocytes (specific leading edge genes from ’reactome interferon signaling’, ’GO IL6 PRODUCTION’, ’reactome IL4 and IL13 signaling’, ’HALLMARK inflammatory response’, ’KEGG JAK STAT signaling’) for branch dependent differential expression using the *BEAM* function from monocle. Select genes were highlighted and categorized based on their expression dynamics along real time and pseudotime.

### Cell frequency analysis

Cell frequencies of activated monocytes gated as HLA-DR+ cells were computed as a fraction of total CD45+CD14+ classical monocytes using flow cytometry data^6^. These cell frequencies were compared across subjects (high vs. low responders) at baseline using a two sided Wilcoxon rank test. The kinetic change of the cell frequency following vaccination was modeled using a mixed effects model with a single random effect for subject ID similar to the models described above. The kinetics over time were modeled using an interaction for time and antibody response group (high vs. low AdjMFC). This interaction model was compared to a timepoint only without the group interaction effect with analysis of variance. The baseline versus day 1 effects for each antibody response group was calculated using the emmeans package.

### Analysis of phospho-signaling responses after stimulation of high and low responder baseline PBMCs using CyTOF

Samples were thawed in a 37°C water bath and washed twice with warmed complete media with Universal Nuclease (Pierce) added. Cells were then washed a final time and resuspended in complete media. 1 million cells per condition were added to individual wells and rested in a tissue culture incubator for 2 hours (37°C, 5% CO_2_). Samples were then stimulated with either PMA/Ionomycin (final concentration [10 ng/mL])/([1μg/mL]); Sigma-Aldrich), LPS (final concentration [1μg/mL]; Sigma-Aldrich), IFN-a (final concentration [10,000U/ml], PBL Assay Science), or left unstimulated. After 15 minutes at 37°C, samples were fixed with paraformaldehyde (2.2% PFA final concentration) for 10 minutes at 25°C. Samples were washed twice with Maxpar Barcode Perm Buffer (1X concentration; Standard Biotools). Samples were then barcoded with Cell-ID 20-Plex Pd Barcoding Kit (Standard Biotools) and incubated at 25°C for 30 minutes. Samples were then washed twice with Maxpar Cell Staining Buffer (Standard Biotools) and combined into corresponding barcoded batches of 5 samples (4 conditions per sample) and washed a final time with Maxpar Cell Staining Buffer. Samples were then stained with a titrated antibody-panel for extracellular markers (Supplementary Table) for 30 minutes at 25°C. After staining, the cells were washed twice with Maxpar Cell Staining Buffer and permeabilized in methanol (Fisher Scientific) overnight at -80°C. The next day, samples were washed twice with Maxpar Cell Staining Buffer, and stained with a titrated panel of antibodies for intracellular signaling markers (Supplementary Table) at 25°C for 30 minutes. Samples were then washed twice with Maxpar Cell Staining Buffer, and labeled with Cell-ID Intercalator Ir ([1:2000] in Maxpar Fix-Perm Buffer; Standard Biotools) overnight at 4°C. The following day, samples were washed twice with Maxpar Cell Staining Buffer and resuspended in 500μL freezing media (90% FBS (Atlanta Biologicals) + 10% DMSO (Sigma-Aldrich)), and stored at -80°C until acquisition. The day of acquisition, samples were thawed and washed twice with Maxpar Cell Staining Buffer and then once with Cell Acquisition Solution (Standard Biotools) before being resuspended in Cell Acquisition Solution supplemented with 10% EQ Four Element Calibration Beads at a concentration of 6 x 10^5^ cells/mL (to approximate 300 events/sec). Samples were acquired on the Helios system (Standard Biotools) using a WB Injector (Standard Biotools). After acquisition, samples were normalized and debarcoded using the CyTOF Software’s debarcoder and normalization tools (Standard Biotools). The panel and protocol were adapted for use at CHI from the Stanford HIMC^110^. The phosphor markers driving the stimulated phenotype and responding cells were automatically defined using the HDStIM R package^70^. The median phosphorylation protein intensity for each individual sample and cell type and stimulation was calculated and modeled with a mixed effects model adjusting or batch and using a random effect for donor ID. The difference in fold change between unstimulated and stimulated cells was calculated using a custom contrast with the emmeans package. CyTOF antibody information is provided in **Supplemental Table 6.**

### Code availability

Code to replicate all analysis in this paper and create all figures is available in the following repository: https://github.com/NIAID/fsc.

### Data availability

All data can be downloaded from the following repository: 10.5281/zenodo.7365959

## Supporting information

Combined supplemental tables.

## Data Availability

Data produced in the present study are available at: https://github.com/niaid/fsc and upon reasonable request to the authors. Select human subject data may not be available for some subjects because study participants did not provide consent for data deposition.

## Acknowledgements

The authors thank members of the Tsang lab for discussions related to this work. This research was supported by the Intramural Research Program of the National Institute of Allergy and Infectious Diseases (NIAID) and Intramural Programs of the NIH Institutes supporting the Center for Human Immunology. Conceptual figures were created using BioRender. The authors thank Eoin Mckinney, Gosia Trynka, Petter Brodin, Sarah Teichman and Ken Smith for constructive comments on this work.

---

**Figure S1.**
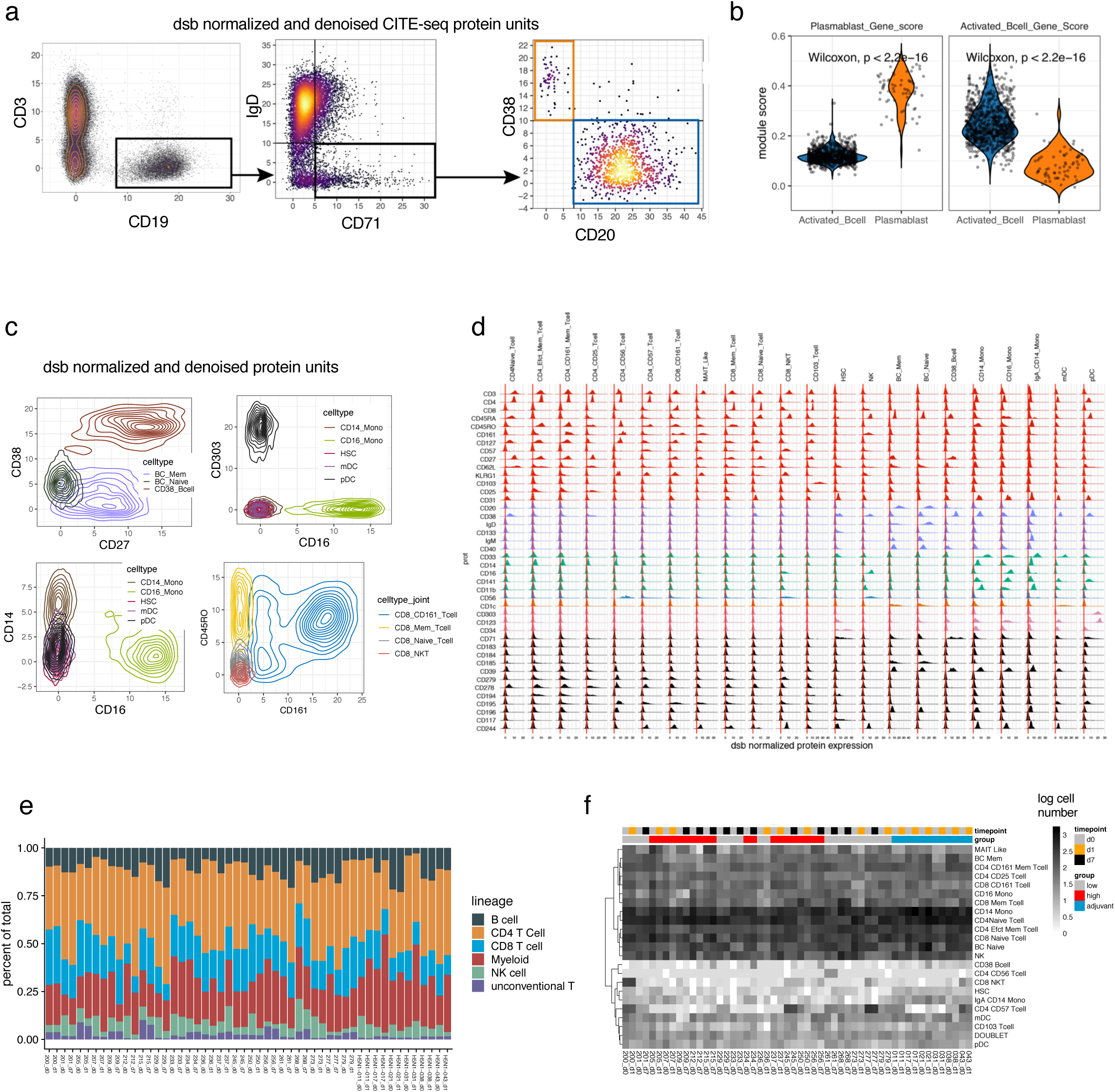
(related to Figure 1) quality control of CITE-seq transcriptome and surface protein phenotypes and clustering. **a.** Manually gated cell populations based on dsb normalized CITE-seq surface protein expression, orange box: plasmablast (CD19+CD71+IgD-CD20-CD38++) and blue box: activated B cell (CD19+CD71+IgD-CD20+CD38+/-). **b.** Transcriptome analysis of gene module scores specific to each gated populations (as in Ellebedy et. al. 2016) p-values shown reflect an unpaired two-sided Wilcoxon test between populations. **c.** Density distribution of dsb normalized protein expression binned by protein based cluster for select populations. **d.** Histograms of dsb normalized protein distribution within each protein-based cluster – rows and columns are hierarchically clustered based on the average expression per cluster. A select subset of proteins are shown and are colored by the main cell populations that they are most informative for discriminating. Red = T cell proteins, light blue = B cell proteins, green = monocyte proteins, dark blue = NK cell proteins, orange = pDC proteins, pink = pDC/HSC markers, black = cell state markers. **e.** The percentage of total cells for each PBMC sample in each major lineage black = B cell, orange = CD4 T cells, blue = CD8 T cells, red = myeloid (all monocytes, HSC, mDC and pDC), green = NK cells, light grey = unconventional T cells (MAIT-like and CD103 + T cells). **f.** The log number of cells per sample by protein based cluster shows that rare individual specific proteins are detected at both timepoints within a given individual.

**Figure S2.**
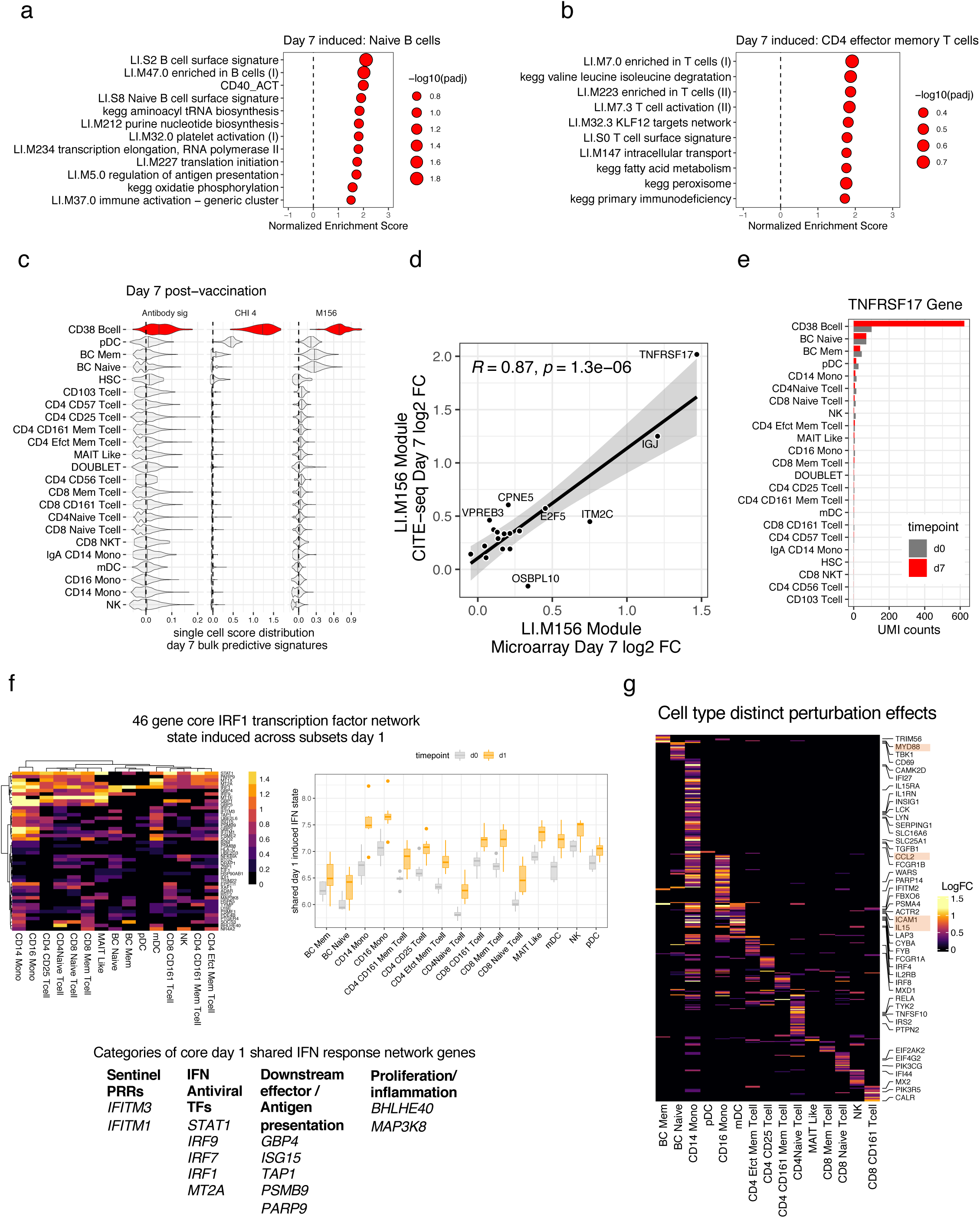
(related to Figure 2). Deconvolution of day 7 antibody titer associated transcriptome signatures and additional shared and cell type specific day 1 cell perturbation phenotypes. **a**. Perturbation phenotypes of naïve B cells day 7 post vaccination. Gene set enrichment as in Fig. 2 based on model adjusted post vaccination effect size, adjusted -log10 p values shown as circle size; pathways with unadjusted p values < 0.01 and NES > 0.1 were included. **b**. As in a, for memory CD4 T cells. **c**. Protein based cell type specificity of day-7 bulk transcriptomic based gene expression signatures predictive of antibody response from previous systems biology studies of influenza vaccination (Supplementary table 1). Single cell level module score distribution shown for day 7 cells for each cell type. **d**. Correlation between genes in M156 detected in CITE-seq (sample level pseudobulk) vs microarray data (Pearson correlation). **e**. Composition of raw counts of the TNFRSF17 gene, a driver of M156 on day 7 across protein-based cell types shows the CD38++ B cells (plasmablasts) are the primary source of the signal. **f**. Left: Heatmap of estimated log fold change 24h post vaccination vs baseline of a core interferon signature shared across subsets – genes selected were increased in at least 5 subsets with logFC > 0.1 and p value < 0.05. Right: the average expression of the core shared interferon signature genes across subsets over time. **g**. Log fold changes as in **f**, here highlighting genes more specifically induced within a single cell type post vaccination.

**Figure S3.**
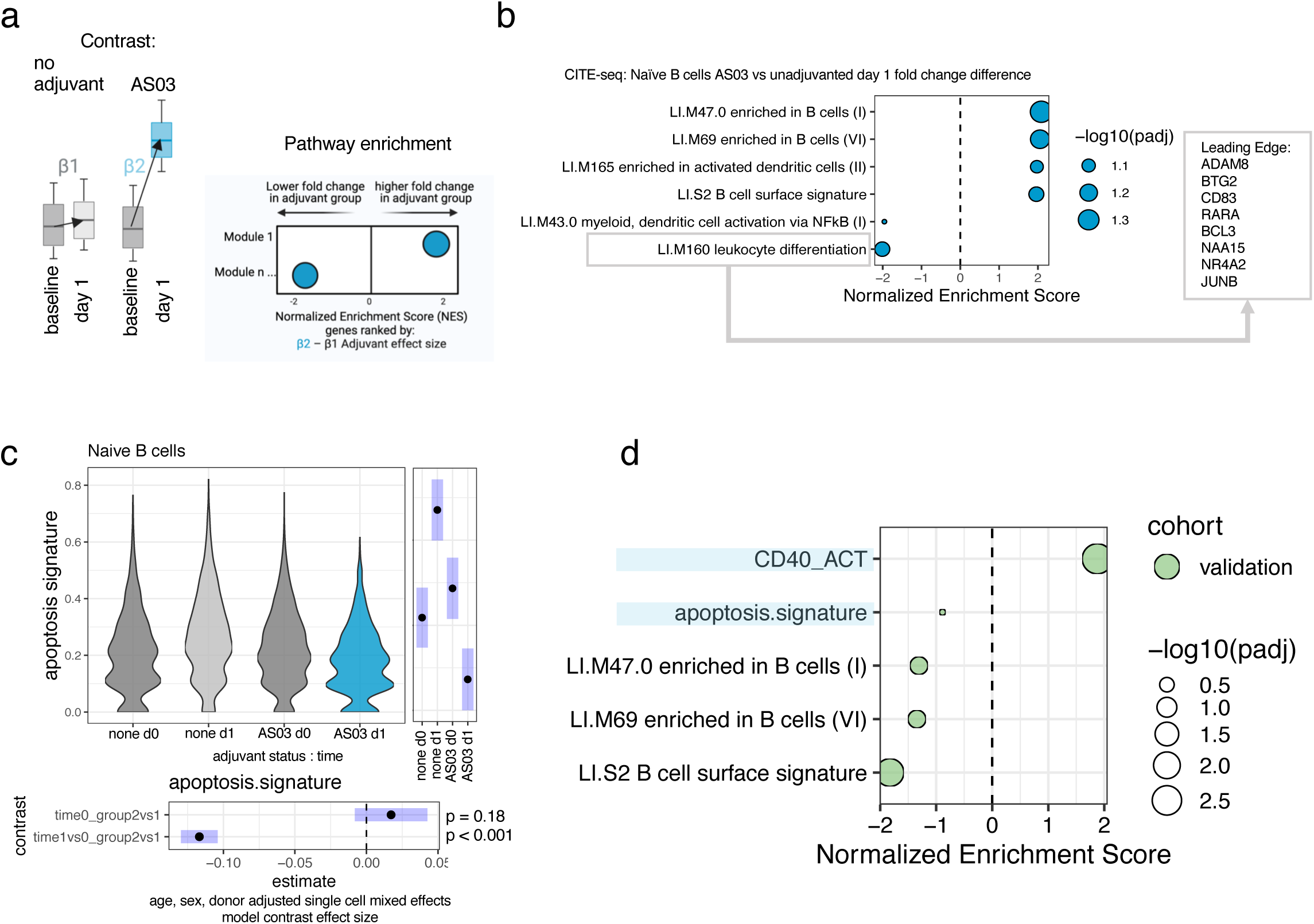
(related to Figure 3) external cohort validation of AS03 perturbation phenotypes and additional analysis of AS03 B cell phenotypes. **a**. Schematic illustrating the contrast applied to each gene comparing unadjuvanted subjects to subjects receiving the AS03 adjuvanted vaccine. The gene set enrichment effect sizes (normalized enrichment score–NES) reflect the genes ranked by the difference in the day 1 fold changes. **b**. Gene set enrichment of the contrast effect shown in a from the mixed effect model for Naïve B cells. **c**. Naïve B cells single cell mixed effects model of a combined apoptosis signature comparing the day 1 fold change AS03 vs unadjuvanted subjects as a function of time post vaccination. The effect size for the time effect for each cohort was opposite, (bottom contrast on bottom margin of plot). The right margin shows the estimated marginal means of the mixed model over levels of the combined vaccine formulation cohort + timepoint variable as calculated by the emmeans package. d. External cohort validation of Naïve B cell CITE-seq derived perturbation phenotypes tested in validation cohort (see Fig 3a) of total sorted B cells (Naïve B cell AS03-specific leading edge genes from CITE-seq analysis as shown in Supplementary Fig 3b tested in all CD19+ cells in the validation cohort). The additional survival signals highlighted in light blue hypothesized to be enriched in naïve B cells after AS03 adjuvanted vaccination based on the M160 genes and top AS03 specific downregulated genes (Fig 3e) and their expression in the CITE-seq cohort include the combined “apoptosis signature” and CD40 activation (CD40 ACT) signature (see methods).

**Figure S4.**
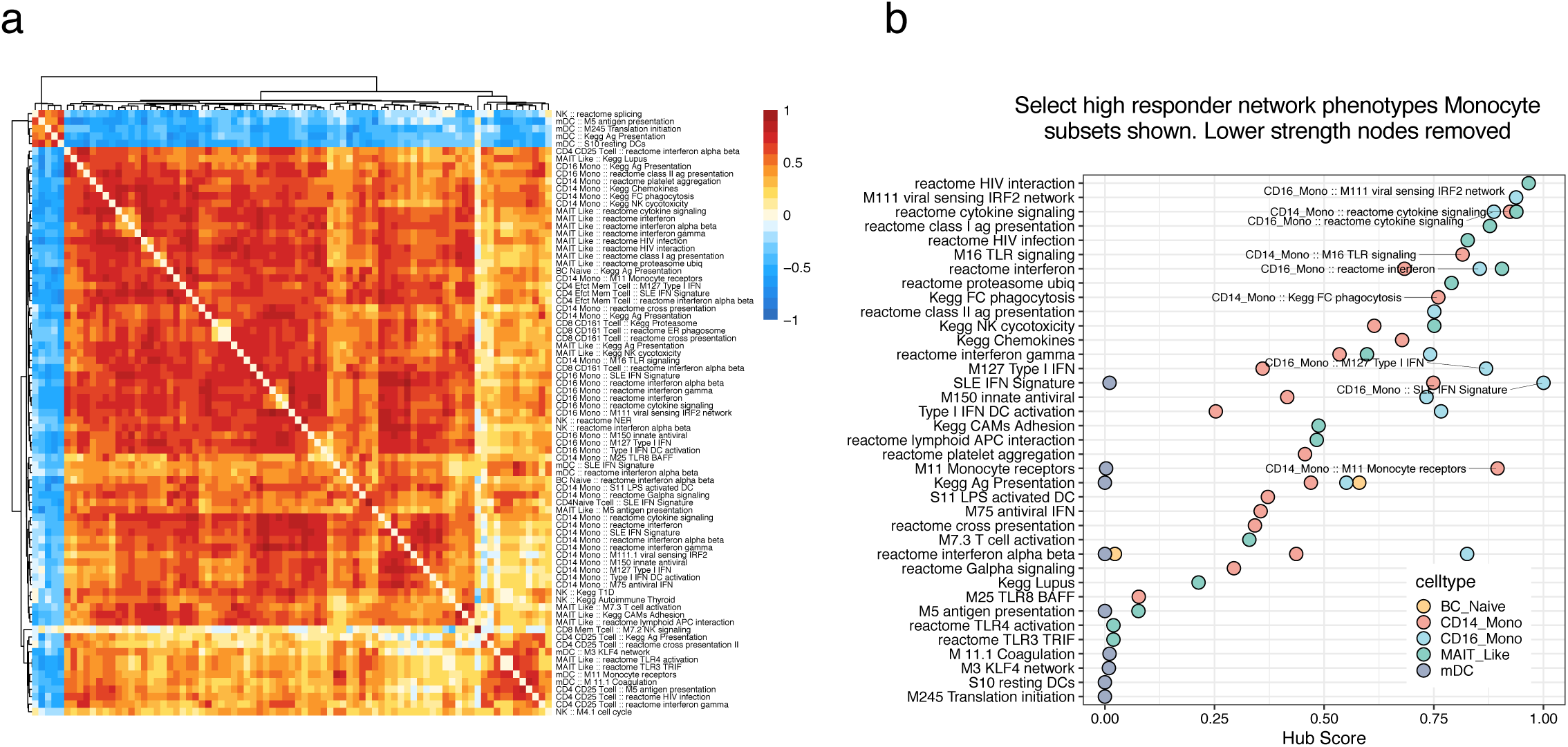
(related to Figure 4) immune setpoint network of high responders. **a.** Correlated multicellular baseline high responder cell phenotypes as a network. The matrix shows the Spearman correlation of expression of each gene module leading edge genes defined in high vs low responders across donors. Correlations within a given cell type are adjusted for gene content (see methods) as exhibited by the diagonal of the matrix (correlations between the same signals) showing a correlation of 0 instead of 1. **b.** The hub score of nodes in the high responder setpoint network after removing edges with correlation adjusted p < 0.05 and those with connection strength (Spearman’s Rho for intercellular connections or shared latent information for intracellular connections) below the median in the network. Nodes highlighted with text include all CD14 and CD16 monocyte nodes including those show in in Figs 4b-c. Points are colored by cell type; the annotation of modules may be the same for a given row (e.g. reactome interferon in CD14 and CD16 monocytes) but the same module is captured by different genes driving the high responder effect in each cell type (e.g. they reflect cell type specific cell phenotypes).

## Notes

### Competing Interest Statement

The authors have declared no competing interest.

### Funding Statement

The work was funded by the Intramural Program of NIAID and the Institutes and Centers supporting the Trans-NIH Center for Human Immunology.

### Author Declarations

Approval provided by NIH Office of Human Subjects Research Protections, Office of IRB Operations.

